# Brazilian Modeling of COVID-19(BRAM-COD): a Bayesian Monte Carlo approach for COVID-19 spread in a limited data set context

**DOI:** 10.1101/2020.04.29.20081174

**Authors:** Samy Dana, Alexandre B. Simas, Bruno A. Filardi, Rodrigo N. Rodriguez, Leandro da Costa Lane Valiengo, Jose Gallucci-Neto

## Abstract

1

**Background:** The new coronavirus respiratory syndrome disease (COVID-19) pandemic has become a major health problem worldwide. Many attempts have been devoted to modeling the dynamics of new infection rates, death rates, and the impact of the disease on health systems and the world economy. Most of these modeling concepts use the Susceptible-Infectious-Susceptible (SIS) and Susceptible-Exposed-Infected-Recovered (SEIR) compartmental models; however, wide imprecise outcomes in forecasting can occur with these models in the context of poor data, low testing levels, and a nonhomogeneous population.

**Objectives:** To predict Brazilian ICU beds demand over time and during COVID-19 pandemic “peak”.

**Methods:** In the present study, we describe a Bayesian COVID-19 model combined with a Hamiltonian Monte Carlo algorithm to forecast quantitative predictions of infections, number of deaths and the demand for critical care beds in the next month in the Brazilian context of scarce data availability. We also estimated COVID-19 spread tendency in the state of São Paulo and forecasted the demand for critical care beds, as São Paulo is the epicenter of the Latin America pandemic.

**Results:** Our model estimated that the number of infected individuals would be approximately 6.5 million (median) on April 25, 2020, and would reach 16 to 17 million (median) by the end of August 2020 in Brazil. The probability that an infected individual requires ICU-level care in Brazil is 0.5833%. Our model suggests that the current level of mitigation seen in São Paulo is sufficient to reach *R_t_ <* 1, thus attaining a “peak” in the short term. In São Paulo state, the total number of deaths is estimated to be around 9,000 (median) with the 2.5% quantile being 6,600 deaths and the 97.5% quantile being around 13,350 deaths. Also, São Paulo will not attain its maximum capacity of ICU beds if the current trend persists over the long term.

**Conclusions:** The COVID-19 pandemic should peak in Brazil between May 8 and May 20, 2020 with a fatality rate lower than that suggested in the literature. The northern and northeastern regions of Brazil will suffer from a lack of available ICU beds, the southern and central-western regions appear to have sufficient ICU beds, finally, the southeastern region seems to have enough ICU beds only if it shares private beds with the publicly funded Unified Health System (SUS). The model predicts that, if the current policies and population behavior are maintained throughout the forecasted period, by the end of August 2020, Brazil will have around 7.6% to 8.2% of its population immune to COVID-19.

## 2 INTRODUCTION

A new unknown-etiology pneumonia was described in Wuhan, China, at the end of 2019. From December 31, 2019, through January 3, 2020, Chinese authorities detected a total of 44 cases of this disease [1]. Over a short period of time, the disease has spread to American and European territories with exponential growth, affecting more than 2 million individuals in 185 different countries/regions [2]. Brazil, the most populous country in Latin America with more than 204.5 million habitants, is the most affected by COVID-19 pandemic. It has a publicly funded Unified Heath System that is responsible for the care of 70 percent of Brazil’s population. The number of critical care beds in Brazil, although not uniformly distributed, is about 28.2 per 100,000 inhabitants [3], comparable to Germany and superior to Italy and Spain. COVID-19 modeling poses a real challenge to epidemiologists and statisticians in developing countries, since most data are underreported and unreliable. Furthermore, Brazil has one of the lowest testing rates per thousand inhabitants (< 1.5 tests/1,000 pop.) in the world [4] and a high probability of widespread underreported cases of COVID-19. Modeling COVID-19 is relevant and necessary, not only to estimate pandemics dynamics but also to forecast health care demands to guide short-term interventions by health authorities and to estimate the impact of social distancing measures. Since March 22, 2020, São Paulo has been under a partial lockdown with social mobility reduced to rates around 50% [5]. In the present study, we describe a probabilistic model for COVID-19 that relies on prior probability distributions over parameters that could fit the complex Brazilian demographics and the scarce data available, and we use this model to estimate the tendency of COVID-19 spread in Brazil, the timing of peak cases, the number of deaths in the country and in the state of São Paulo, and the stress of the COVID-19 peak on the ICUs in the Brazil health system.

## 3 METHODS

### 3.1 Imputed variables

The serial interval is one of the most important variables in epidemiological studies of epidemics. It can be defined as the time between the onset of the disease in the primary case (transmitter) and the onset of the disease in the secondary case (infected by the primary case) [6], and it is the fundamental variable used to estimate transmission capacity and turnover for generating new cases [7], *R*_0_, or the basic reproductive number, represents the average number of people who will be infected from a sick patient [8]. Curiously, data for these two variables from the literature followed two patterns. Data from the beginning of the COVID-19 epidemic suggested a longer serial interval, of about 7.5 days [9], which was the same order of magnitude as that of SARS (8.4 days) [10]. However, all articles and publications subsequent to this phase in early 2020 found shorter intervals, which is consistent with the practical evolution of the disease in practically all affected countries. Therefore, we chose to incorporate the most current data compatible with reality into the model and to disregard the findings from the beginning of the epidemic.

Conceptually, when the serial interval number is less than the estimated disease incubation period, in practice the transmission by presymptomatic patients is more frequent. As it is known that a large portion of infected people can be asymptomatic [11] and, therefore, silent transmitters of the disease, isolation of only symptomatic patients is an ineffective strategy for controlling the epidemic; thus, social isolation is mandatory.

Using Nishiura et al. [7] as a basis, we take the median serial range to be 4 and the mean serial range to be 4.7. These values are consistent with those found by other researchers [12, 13]. *R*_0_ values in the literature vary between 2.2 and 6.47 [9, 14, 15, 16, 17]. We chose to adopt the values found by Liu et al [18] in a systematic literature review: an average *R*_0_ of 3.28 and a median *R*_0_ of 2.71. It is known that the virus transmission capacity depends on the viral load, and it is also possible that the virulence of the disease is related to the amount of viral particles inhaled over time [19]. Furthermore, seriously ill patients continue to transmit the disease indefinitely, even after the presence of IgG in the blood [20]. Our modeling efforts try to incorporate these different scenarios.

We adopted the following premises: (1) Asymptomatic patients transmit the virus for a period between 6 and 10 days; oligosymptomatic patients transmit the virus for up to 14 days [9]; and seriously ill patients transmit for up to 4 weeks, but we consider 10 days for these patients as they will be isolated in an ICU. (2) The virus incubation time is the variable with the least variability among authors: 5.2 days according to Li et al. [9], 5.1 days according to Zhang et al.,[13] and 3 to 7.2 days according to Yang et al. [21] (3) Of the main risk factors, without a doubt, the most clinically important is age. Mortality after 62 to 65 years becomes extremely significant [22, 23], with about 90% of deaths occurring in this age group. In a recent systematic review of the literature by Ioannidis et al. [22], fewer than 10% of those who died were under the age of 65, and fewer than 1% percent of deaths occurred in those under 40. Only 4%–9%: of deaths occurred in those who are between the ages of 40 and 65. (4) Men are 2 to 3 times more likely to die. (5) Hypertension [23] and obesity [24] are significant risk factors, among others [25].

Due to these factors and the known presence of a large number of asymptomatic people seen in countries whose public health policies have facilitated the testing of a large portion of the population [26], we added to the model a correction factor that modifies the chance of death as described by Ioannidis et al. [22]. In this correction factor, we consider a gradual change in the number of asymptomatic patients that is in inverse proportion to their age.

Regarding the timing of disease progression, we consider the time from onset of symptoms to admission to ICU to be 3 to 7 days, and the time between onset of symptoms and death to be about 3 weeks. We assume that 9% of symptomatic cases need admission to the ICU and that 25% of those ICU patients die (Grasselli et al. [23]). We also add that the average age of patients admitted to the ICU is similar to the average age of people who died (64 years old), and the median of the Kaplan-Meier curve is 25 days for patients admitted to the ICU [21].

Other factors that can be extremely relevant but lack proof of cause and effect as they are eminently associative are seasonality [27] and the Bacillus Calmette-Guerin (BCG) vaccine [28, 29]. BCG is a vaccine used against tuberculosis, and according to the distribution of this disease, the countries that mostly use the vaccine are developing countries like Brazil. There is an important association, of the order of ten times, between vaccination coverage and number of fatal cases by COVID-19 [28, 29]. However, the data are associative and the level of evidence is weak; thus, it is impossible to establish a causal relationship. Considering that there is a biological plausibility that justifies partial coverage against coronavirus by an indirect effect, and given the magnitude of the finding, we chose to include it in the model, giving this vaccine the benefit of the doubt. If it does not have a protective effect, the pessimistic scenario does not change, but the lack of effect will be observed when examining the optimistic scenario.

The effects of climate and temperature could also affect the transmission capacity of the virus, because the viability of the virus seems to be influenced by sunlight, temperature, and humidity. Similarly, it is extremely common to have seasonal characteristics in the incidence of colds, and the coronavirus is influenced by season [27]. One of the reasons that we may not have seen the expected intensity of the epidemic in our country may be the fact that transmissibility is affected by these seasonal issues. The country’s first virus identification was in February, in the middle of summer. If the seasonal factor is truly significant, the worst months could be June and July. In contrast, we find that the values fed in the model (serial range and *R*_0_) come from countries that were in theoretically favorable seasonality.

We hypothesize, therefore, that if seasonality is of great importance in our environment, there will be no worsening beyond the more pessimistic scenario. Otherwise, there would be a non-accentuated increase in the mortality curve.

### 3.2 Model description

We build our model upon the model introduced in Flaxman et al [30].

Let us fix some notation that will be used throughout the manuscript. We denote by *R* the region of interest, such as São Paulo State, or the entire Brazil country. The parameters of the gamma distributions will always be shape and scale, and the parameters of the lognormal distributions will always be log-mean and log-standard deviation.

Let *M_t_,_R_* be the number of deaths in the region *R* at the tth day. We assume that

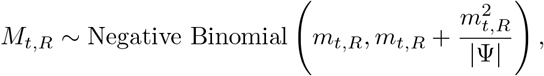

where *m_t_,_R_* is the expected number of deaths and we assume that *ψ* ~ *N*(0, 1).

We are assuming the Negative Binomial distribution for simplicity. It assumes an infinite population, but we ensure the existence of herd immunity by using the factor (4). One could also adapt the above construction to a Beta Binomial distribution, which has assumes a finite population, but the results will probably be similar since the population and the probability of living are large and the Beta Binomial distribution can be approximated by the Negative Binomial for large population and large probability of living.

At this point, our strategy differs from that seen in [30]. Indeed, we will model the expected number of deaths at the *t*th day and region *R* by modelling the expected number of individuals infected by COVID-19 that will need ICU-level care.

To this end, let *N_t_,_R_* be the expected number of individuals infected by COVID-19 that will need ICU-level care at the *t*th day on the region *R*. Then, we consider the following structure:

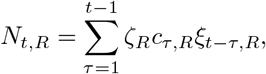

where *ζ_R_* is the probability that a person will need ICU-level care at region *R*, *C*_τ_,*_R_* is the number of infected individuals at time *τ* in the region *R* (which will be modelled below) and *ξ_t−τ_,_R_* is the conditional probability of a person that got infected in the day *τ* will need ICU-level care at the day *t*, conditional to the fact that the individual will, indeed, need ICU-level care.

By considering the infection-fatality-rate given in [31] and the results of [32], we arrived at the following approximation:

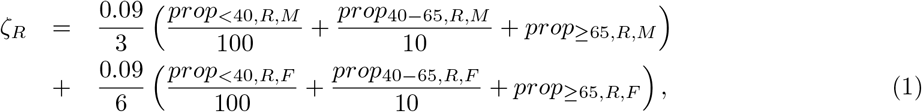

where *prop_<_*_40_*_,R,M_* (*prop_<_*_40_*_,R,F_*) stands for the proportion of male (female) individuals at region *R* whose age are less than 40, *prop*_40−65_*,_R_,_M_* (*prop*_40−65_*_,R,F_*) stands for the proportion of male (female) individuals at region *R* whose age are greater or equal than 40 and less than 65, and *prop_≥_*_65_*_,R,M_* (*prop_≥_*_65_*_,R,F_*) stands for the proportion of male (female) individuals at region *R* whose age are greater or equal than 65.

We will now describe *ξ_s_,_R_*, which is the conditional probability of an individual in region *R* that will need ICU-level care infected at day 0 need the ICU care at day s. We will use two auxiliary random variables to this end. Let *T_inc_* be the incubation time of COVID-19 and *T_ICU_* be the duration between a pacient, that will need ICU-level care, be infected and need the ICU care. From [33], we will assume that

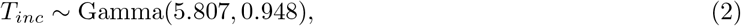

and from [34], we will assume

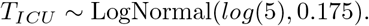

Then, we will let

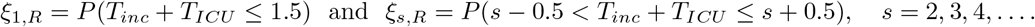

We follow the dynamics of *C_τ_,_R_* used in [30]. Thus, we have

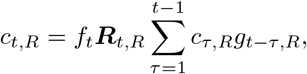

where *f_t_* is a factor to ensure the existence of herd immunity and is given by (4), ***R****_t_,_R_* is the reproduction number at time *t*, that is, it represents the average number of secondary cases generated by a primary infection at day *t*, and *g_t−τ_,_R_* is the probability that an individual infected at time *τ* begins infecting at time t. We will describe this probability distribution given in [30]. By [13] and [35] we may conclude that there is evidence of presymptomatic transmission. The time distribution given in [30] is given by

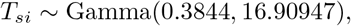

which has mean 6.5 and coefficient of variation of 0.62. This seems to take into account the presymptomatic transmission as well as the reported incubation times. This choice also provided good forecast perfomance on empirical data.

We now describe our estimation of the reproduction number ***R****_t_,_R_*. This is a crucial step and we provide a suggestion that we believe can be improved in the future.

Let *t\** be the first day in which the number of reported cases is greater than 1 and *t\*\** the first day in which the number of reported deaths is greater than 1. We let

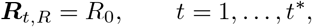

where

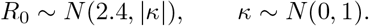

For *t* = *t\** + 1,…,*t\*\** we use the history of reported cases to estimate *R_t_* following the Bayesian approach given in [36], whose details can be found in their Appendix within the supplementary data. This approach is implemented in the function *estimate_R* in the package `EpiEstim` for the `R` statistical software. During this period, even though the number of reported cases is not very reliable due to undernotifications, its trend is useful and that is what we are using. For *t* = *t\*\** +1,… we use the reported deaths to estimate *R_t_* following the same approach we used for the reported cases. We expect the reported deaths to be more reliable, but the figures are usually very similar. We considered windows varying from 1 to 3 weeks for calibration. Typically, the best choice was among the window with 2 weeks, or an average between the estimated ***R****_t_,_R_* between 2 and 3 weeks. Shorter windows obtain instantaneous changes, but suffers from noise, longer windows are smoother and capture longer trends, but may miss changes in trends. The balance among the longer and shorter windows is achieved through calibration. Thus, by applying the method described in [36] we obtain 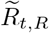. We define

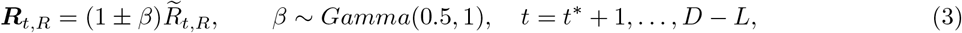

where the sign (plus or minus) should be chosen in order to compensate a prior belief that a model is underestimating or overestimating *R_t_*, *D* is the total number of days and *L* is the chosen lag. To estimate ***R****_t,R_* for the remaining, out-of-sample, days needed in the forecast, we estimate (by least squares) *λ*, *γ* and *δ* (they are not estimated within the Bayesian procedure, they are obtained separately) such that, 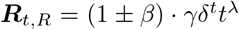. This choice has ensured good forecast behavior in the datasets we considered. It is also indirectly calibrated by *β*.

It is very important to notice that this choice also assumes that a trend found in the reproduction number in the past will be maintained in the future, so it is very sensitive to changes in public policies, public behavior and/or medical advances. Therefore, in an event such as changing isolation policies, the forecast is compromised. Finally, in order to simulate such an event we include covariates in the structure of the reproduction number to indicate the occurence of the event.

We will now model, at the same time, the occupation of ICU beds and the number of deceased individuals due to COVID-19. Recall that *N_t_,_R_* is the number of individuals that will need ICU-level care. Let *TB_R_* be the total number of ICU beds in the region *R*. We will denote by *AB_t,R_* the number of available beds in the *t*th day on the region *R*. We are initially assuming that the occupation rate for other reasons is constant through time. One can easily adapt to change the occupation through time, for instance, due to changes in government policies.

We will need the following variables: *T_dec,ICU_ ~* Gamma(3.15,0.28), the conditional duration between entering the ICU and deceasing, conditional on the event that the individual will die; *T_live,ICU_ ~* Gamma(16.33,1.16), the conditional distribution between entering the ICU and being released, conditional on the event that the individual will live; *D ~* Beta(2, 5), the probability that an individual that enters the ICU will die. If an individual needs ICU-level care and there is no bed available, then this individual will necessarily die *T_dec,non-ICU_ ~* LogNormal(log(2), 0.3) days later.

We now proceed as follows. Initialize *m_t_,_R_* = 0 for each day *t*. Let *τ*_1_ = min{*t*; *N_t_,_R_ >* 0}. For *t* = 1,…, τ_1_ − 1, *AB_t,R_* = *TB_t,R_*. On time *τ*_1_, we have

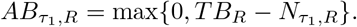

Then, update the numbers of deceased at day *τ*_1_ + *T_dec_,_ICU_*:

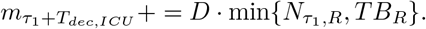

We have to take into account the individuals that will not have bed available:

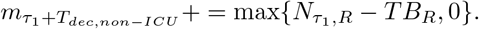

We will now occupy the beds of ICU. Consider a cutoff on the number of maximum days (for computational enhancement), say *C_days_*. We considered as cutoff day such that with probability 95% that will die, will already be dead by that day, namely, we considered *C_days_* = 24 days. But, this parameter may be chosen at will. We also truncate *T_dec,ICU_* and *T_dec,non−ICU_* so that they will always be less than or equal to *C_days_*.

Define the 2-dimensional vectors 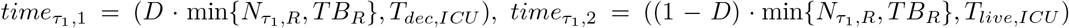, and for *i* = 3,…,*C_days_* + 1 let 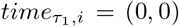. Thus, we have *C_days_* + 1 vectors *time*, one for each possible duration in the ICU, and one more, to ensure the consistency of the algorithm, that is, if the algorithm says that there are available beds, then we will find the coordinates of the empty beds (among *i* = 1,…,*C_days_* + 1) accordingly. Finally, let the number of survivors at day *τ*_1_ (even though it is not known by the physician at this point) be given by

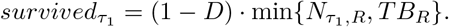

For the subsequent days *t > τ*_1_, proceed as follows. For each *i* = 1,…,*C_days_*, let *time_t,i,_*_2_ be the second coordinate of *time_t,i_*, and do *time_t,i,_*_2_ = max{time*_t−_*_1,_*_i_*,_2_ − 1,0}, to indicate that one day have passed. Now, count the number of available beds: *AB_t,R_* = #{*i*; *time_t_,_i_*,_2_ *<* 1}. Now, occupy the beds:

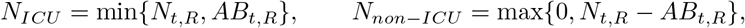

where *N_ICU_* is the number of individuals that occupied beds in the ICU and *N_non−ICU_* is the number of individuals that could not find an available bed. Let *i*_1_ and *i*_2_ be two vacant positions of beds, which is guaranteed from the fact that we chose *C_days_* + 1 slots, with maximum staying period at ICU of *C_days_*. Hence, let 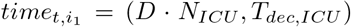 and 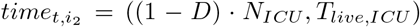. Also, update the expected number of dead individuals accordingly:

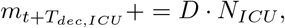

and

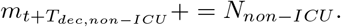

The number of survivors at day *t* is given by

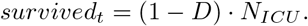

This concludes the modelling of deceased. We now have to take into account the people that were immunized, so we can compute the herd immunity factor.

Initialize *f*_1_ = 1. On the tth day, we will consider the infected individuals on the day *t* − 1 (we cannot use the individuals of the *t*th day, since it depends on *f_t_*, which is not yet computed) that will never go to the ICU:

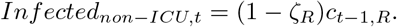

Therefore, we defined the immunized individuals up to time *t* as

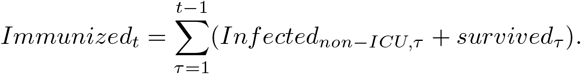

Let *prop_herd_* be a proportion in which a population is considered to enjoy the herd immunity. Typical numbers vary from 0.5 to 0.75. We used *prop_herd_* = 0.6. Then, the herd immunity factor is given by

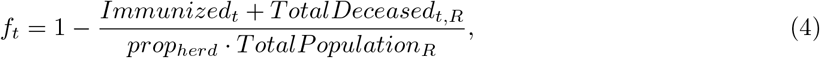

where *TotalPopulation_R_* stands for the total population of the region *R* and

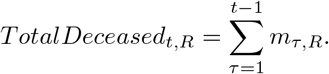

As in [30], we also assume that the seeding of new infections begins 30 days before the day after a region has cumulatively observed 10 deaths, and we also seed our model with 6 sequential days of infections drawn from *c*_1,_*_R_*,…,*c*_6,*R*_ ~ *Exponential*(*τ*), where *τ* ~ *Exponential*(0.03).

We fitted the model in the real data using the statistical softwares `R` (see [37]) and `Stan` (see [38]) through an adaptive Hamiltonian Monte Carlo (HMC) sampler. We ran 4 chains for 4000 iterations with 2000 iterations of warmup and a thinning factor of 4 to obtain 2000 posterior samples. Posterior convergence was assessed using the Rhat statistic.

### 3.3 ICU Beds From the Public and Private Sectors

In Brazil, the ICU beds are unevenly distributed between the public and private sectors. We will now model a situation in which people that do not have health insurance plan are not able to use ICU beds from the private sector, but the people who have health insurance plans may use ICU beds from both the private and public sectors, but try first to use the ICU beds from the private sector.

This is a very important and realistic situation to model. Indeed, most of the population do not have health insurance plans. Furthermore, the number of ICU beds per 100 thousand inhabitants vary a lot throughout the states of Brazil.

We will now describe how we modified the previous algorithm to model this situation. We begin by obtaining a list of health insurance coverage for each state of Brazil. This data is available at the home page of the National Regulatory Agency for Private Health Insurance and Plans from Brazil. With this data, we are able to compute the proportion of people in each region *R* that have health insurance plan, say *prop_health−plan,R_*.

Finally, we obtain the number of ICU beds from the public and private sectors for each state of Brazil. This list is obtained in the CNES-DATASUS database. Let *TB_public,R_* and *TB_private,R_* be the numbers of ICU beds from the public and private sectors of region *R*, respectively.

The modification goes as follows: Initialize *m_t_,_R_* = 0 for each day *t*. Let *τ*_1_ = min{*t*; *N_t_,_R_ >* 0}. For *t* = 1,…,*τ*_1_ − 1, *AB_t,public,R_* = *TB_public,R_* and *AB_t,private,R_* = *TB_private,R_*. On time *τ*_1_, we have

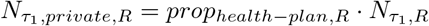

and

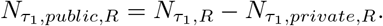

Thus,

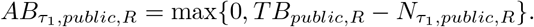

Then, update the numbers of deceased at day *τ*_1_ + *T_dec_,_ICU_*:

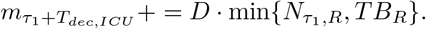

We have to take into account the individuals that will not have bed available:

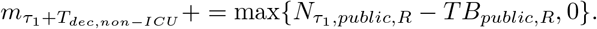

We will now occupy the beds of ICU from the public sector. As before, we will also consider a cutoff on the number of maximum days (for computational enhancement), say *C_days_*.

Define the 2-dimensional vectors 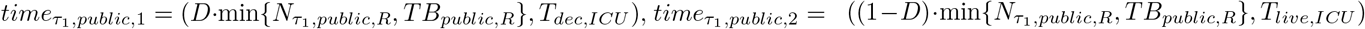, and for *i* = 3,…,*C_days_* +1 let 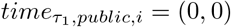. Thus, we have *C_days_* + 1 vectors *time*, one for each possible duration in the ICU, and one more, to ensure the consistency of the algorithm, that is, if the algorithm says that there are available beds, then we will find the coordinates of the empty beds (among *i =* 1,…,*C_days_* + 1) accordingly. Finally, let the number of survivors at day *τ*_1_ (even though it is not known by the physician at this point) be given by

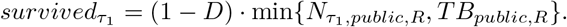

Now, we will move to the ICU beds from the private sector. Recall that *τ*_1_ = min{*t*; *N_t_,_R_ >* 0}. We have

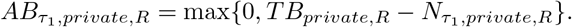

Recall that we are assuming that a person from the private sector may use available beds from the public sector. Thus, compute the number of available beds from the public sector after the previous occupation:

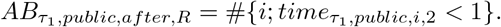

Then, update the numbers of deceased at day *τ*_1_ + *T_dec,ICU_*:

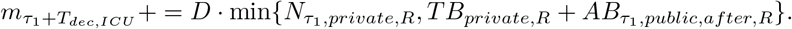

We have to take into account the individuals that will not have bed available:

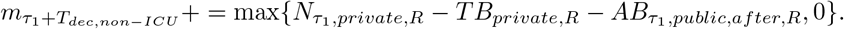

We will now occupy the beds of ICU from the private sector. As before, we will also consider a cutoff on the number of maximum days (for computational enhancement), say *C_days_*.

Define the 2-dimensional vectors 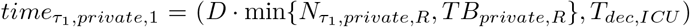, 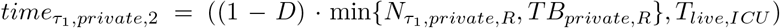, and for *i* = 3,…,*C_days_* + 1 let 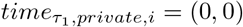. Thus, we have *C_days_* +1 vectors *time*, one for each possible duration in the ICU, and one more, to ensure the consistency of the algorithm, that is, if the algorithm says that there are available beds, then we will find the coordinates of the empty beds (among *i =* 1,…,*C_days_* + 1) accordingly. Finally, let the number of survivors at day *τ*_1_ (even though it is not known by the physician at this point) be given by

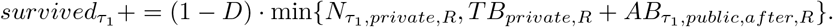

Also, if 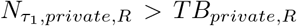, we need to occupy the available public beds accordingly. So, do the following changes: 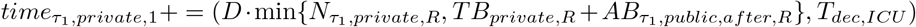 and 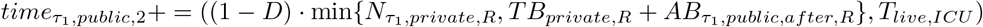.

For the subsequent days *t >* τ_1_, proceed as follows. For each *i =* 1,…,*C_days_*. We will begin by dealing with the ICU beds from the public sectors. Let *time_t_,_public_,_i_*,_2_ be the second coordinate of *time_t,public,_*_1_, and do *time_t_,_public_,_i_*,_2_ = *max{time_t−_*_1_*,_public_,_i_*,_2_ − 1, 0}, to indicate that one day have passed. Now, count the number of available beds: *AB_t_,_public_,_R_* = #{*i*; *time_t_,_public_,_i_*,_2_ *<* 1}. Now, occupy the beds:

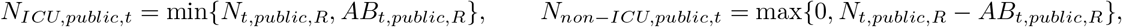

where *N_ICU_,_public,t_* is the number of individuals that occupied beds in the public ICU at time *t* and *N_non−ICU,public,t_* is the number of individuals that could not find an available public ICU bed at time *t*. Let *i*_1_ and *i*_2_ be two vacant positions of beds, which is guaranteed from the fact that we chose *C_days_* + 1 slots, with maximum staying period at ICU of *C_days_*. Hence, let 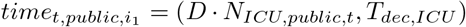 and 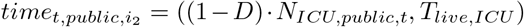. Also, update the expected number of dead individuals accordingly:

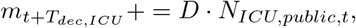

and

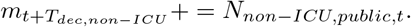

The number of survivors at day *t* is given by

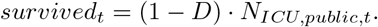

Now, let us deal with the ICU beds from the private sector. Let *time_t_,_private_,_i_*,_2_ be the second coordinate of *time_t_,_private_,_i_*, and do *time_t_,_private_,_i_*,_2_ = max{time*_t_*_−1,_*_private_,_i_*,_2_ − 1,0}, to indicate that one day have passed. Now, count the number of available beds: *AB_t,private,R_* = #{*i*; *time_t,private,i,_*_2_ *<* 1} and *AB_t,public,after,R_* = # {*i*; *time_t,public,i,_*_2_ *<* 1}. Now, occupy the beds:

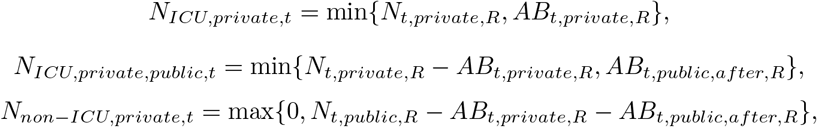

where *N_ICU_,_private,t_* is the number of individuals that occupied ICU beds from the private sector at time *t*, *N_ICU,private,public,t_* is the number of individuals with health insurance that occupied ICU beds from the public sector since ICU beds from the private sector were not available and *N_non−ICU,private,t_* is the number of individuals with health insurance plans that could not find an available public ICU bed at time *t*. Let *j*_1_ and *j*_2_ be two vacant positions of beds, which is guaranteed from the fact that we chose *C_days_* + 1 slots, with maximum staying period at ICU of *C_days_*. Hence, let 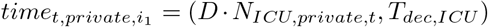 and 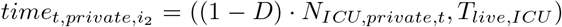. Finally, let *i*_1_ and *i*_2_ be the vacant positions obtained in time *t* from the public ICU beds, and do the following update:

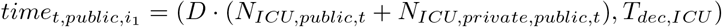

and

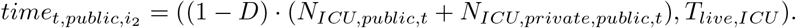

Also, update the expected number of dead individuals accordingly:

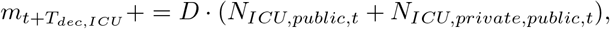

and

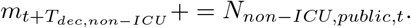

The number of survivors at day *t* is given by

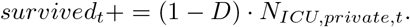

The remaining of the algorithm being the same as before.

## 4 Results

We fitted data from Brazil and São Paulo. The data from Brazil was obtained as a summation of the results across the different regions from Brazil. The different regions from Brazil have heterogeneous age pyramids as well as different ICU capabilities. In Figure 1 we see the age pyramids of Brazil and Italy, respectively. It is noteworthy that the Brazilian age pyramid is much younger than Italy’s pyramid. The source of the brazilian data is https://www.ibge.gov.br/estatisticas/sociais/populacao/9109-projecao-da-populacao.html?=&t=downloads and the italian data is https://population.un.org/wpp/Download/Standard/Population/.

**Figure 1:**
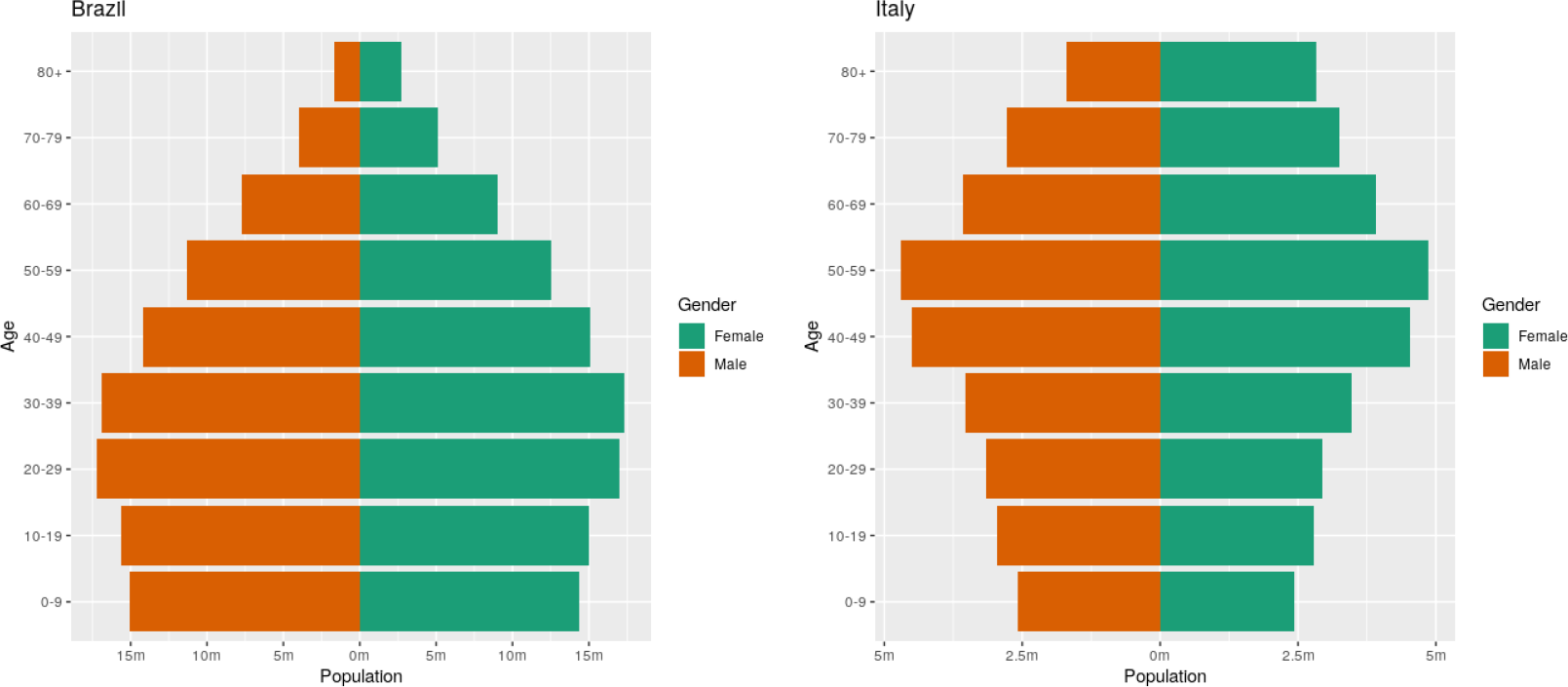
Age pyramids of Brazil and Italy.

In Table 1 we see the estimates of the probability of an infected individual require ICU-level care in Brazil, Italy, and across the different regions from Brazil as well for São Paulo using the formula in equation (1).

**Table 1:** Probability of an infected individual require ICU-level care across different regions.

| Region | ICU probability of an infected individual |
| --- | --- |
| Italy | 1.1965% |
| Brazil | 0.5833% |
| North of Brazil | 0.3968% |
| Northeast of Brazil | 0.5317% |
| Central-West of Brazil | 0.5110% |
| Southeast of Brazil | 0.6424% |
| South of Brazil | 0.6627% |
| São Paulo | 0.6301% |

It is noteworthy that the figures in Table 1 are not far apart to those obtained by combining the numbers in [34] with the age pyramids of across the regions and homogeneous attack rate. It is also remarkable that it becomes explicit in Table 1 the heterogeneous characteristic of the regions of Brazil. This can also be seen in the age pyramids of the different regions of Brazil in Figure 2. We considered the Federal District separately since it differs a lot from the remaining states of the Central-West region.

**Figure 2:**
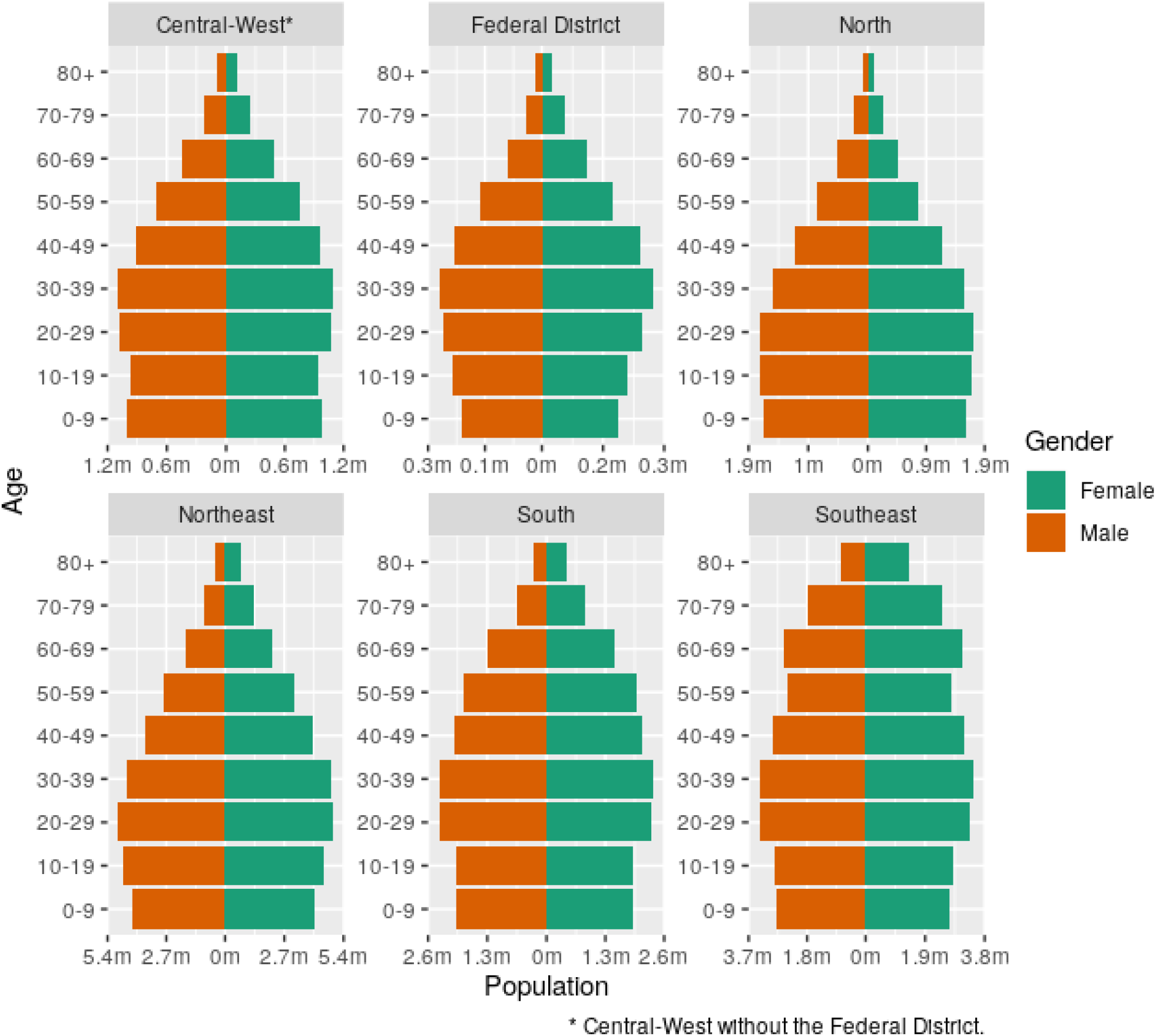
Age pyramids of the regions of Brazil.

Another very important data we need is the number of ICU beds available in Brazil. Figure 3 contains the number of ICU beds per 100 thousand inhabitants in Brazil, Italy and São Paulo, and also in the different regions of Brazil. It is noteworthy that Brazil has several ICU beds, which is due to the high number of severe traffic accidents as well as the high homicide rates. The source of the data is the CNES-DATASUS base from Brazil. For Italy, the source is [39].

**Figure 3:**
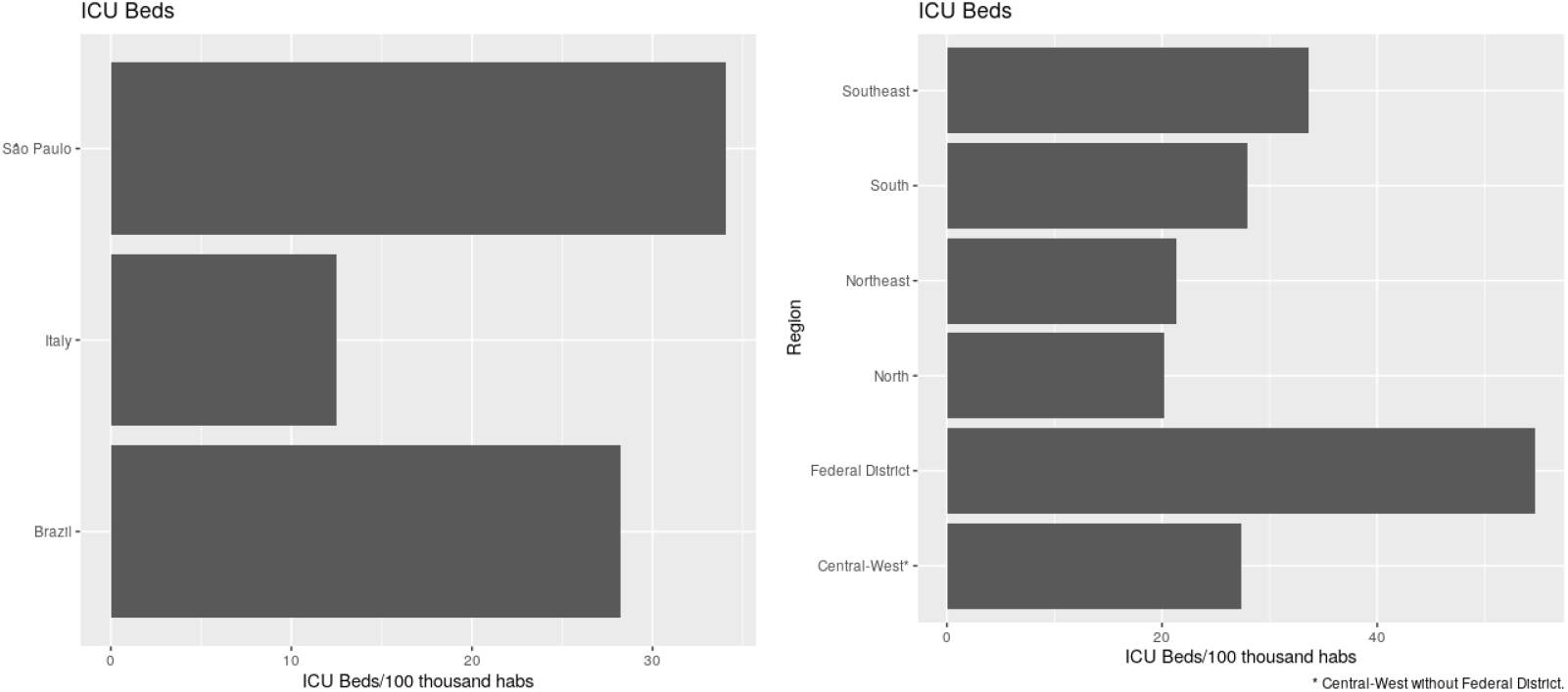
ICU Beds per 100 thousand inhabitants in São Paulo, Brazil and Italy on the left and the different regions of Brazil on the right.

### 4.1 Model fitting in the state of São Paulo

In this subsection we will provide the results of the model fitting to the data of COVID-19 deaths in the state of São Paulo, Brazil. We considered the data available at https://covid.saude.gov.br/ until April 20, 2020.

The model convergence was attained since we obtain a mean of Rhat statistic 1.01 and standard deviation of 0.005.

We manually calibrated the *α* in (3) to account for the occupation of ICU beds currently seen on São Paulo. The result is that the accumulated number of deaths seems to be higher than the observed counterpart, but we also expect this to be true since we believe the number of deaths reported is currently lower than the reality due to latency of testing.

We begin by showing, in Figure 4 the fitted model and the predicted values for the next seven days. The quality of Brazil’s data is very poor. For instance, on mondays, holidays or weekends the reported number of deaths is recurrently very low. Thus, to assess the quality of the model we find it useful to observe the accumulated number of deaths instead of the daily number of deaths. It is also noteworthy that the accumulated number of deaths seems to be decreasing.

**Figure 4:**
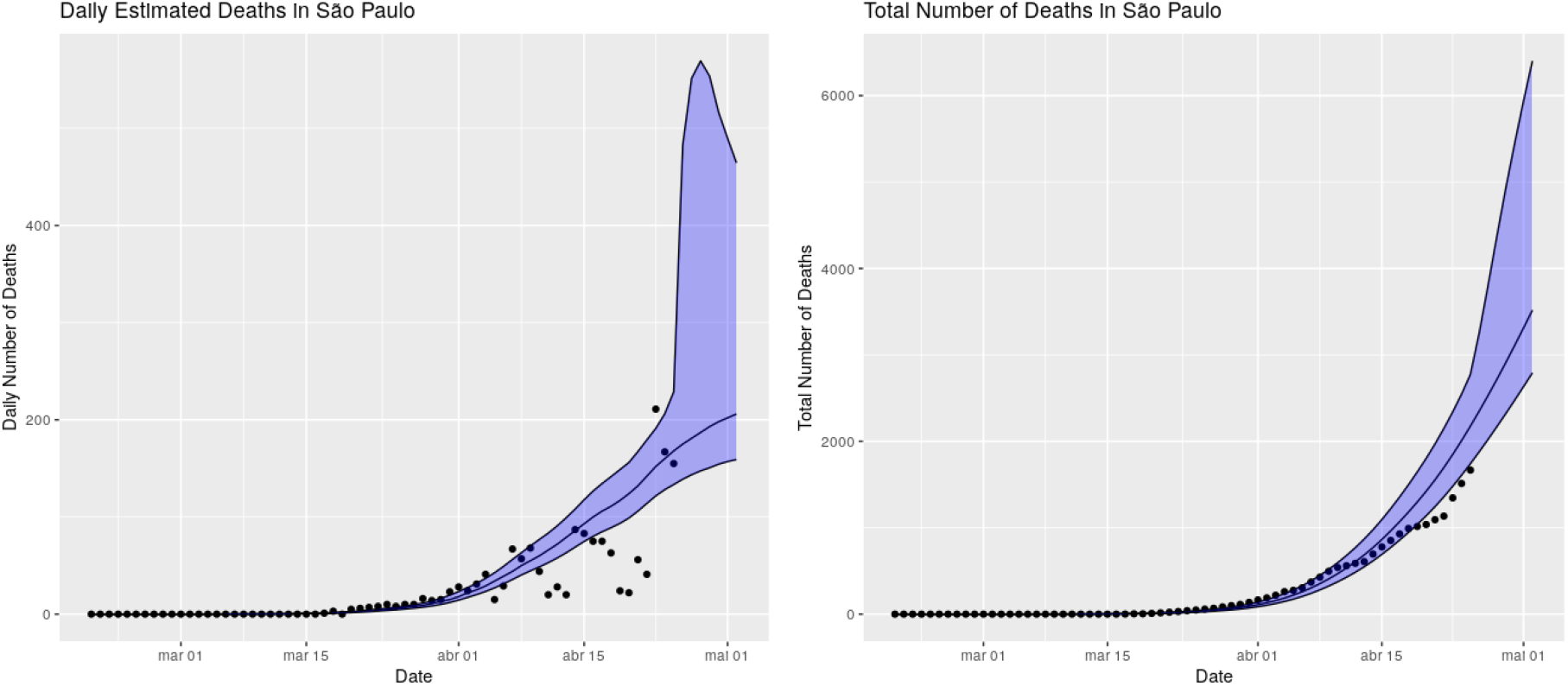
Fitted model and predicted values for the next seven days.

We will now provide our long-term predictions. As it is well-known long-term predictions are unreliable since they assume that the trend seem by the model until today will remain in force through the whole forecast period. Nevertheless, we find they are useful to see to which direction our current policies are leading.

In Figure 5 we provide the predictions of the model until the end of August. The first important remark is that our model suggests that the current level of mitigation seen in São Paulo is enough to reach *R_t_ <* 1, thus attaining a “peak” in the short term. The “peak” is being estimated for the first week of may, 2020. The second observation is that the total number of deaths is being estimated at around 9000 (median) with 2.5% quantile being around 6400 deaths and the 97.5% quantile being around 14400 deaths. These estimated totals vary a lot through the days due to the exponential nature of the model and the bad quality of the data seen in Brazil, but the model has been very consistent with the estimated date of the “peak”.

**Figure 5:**
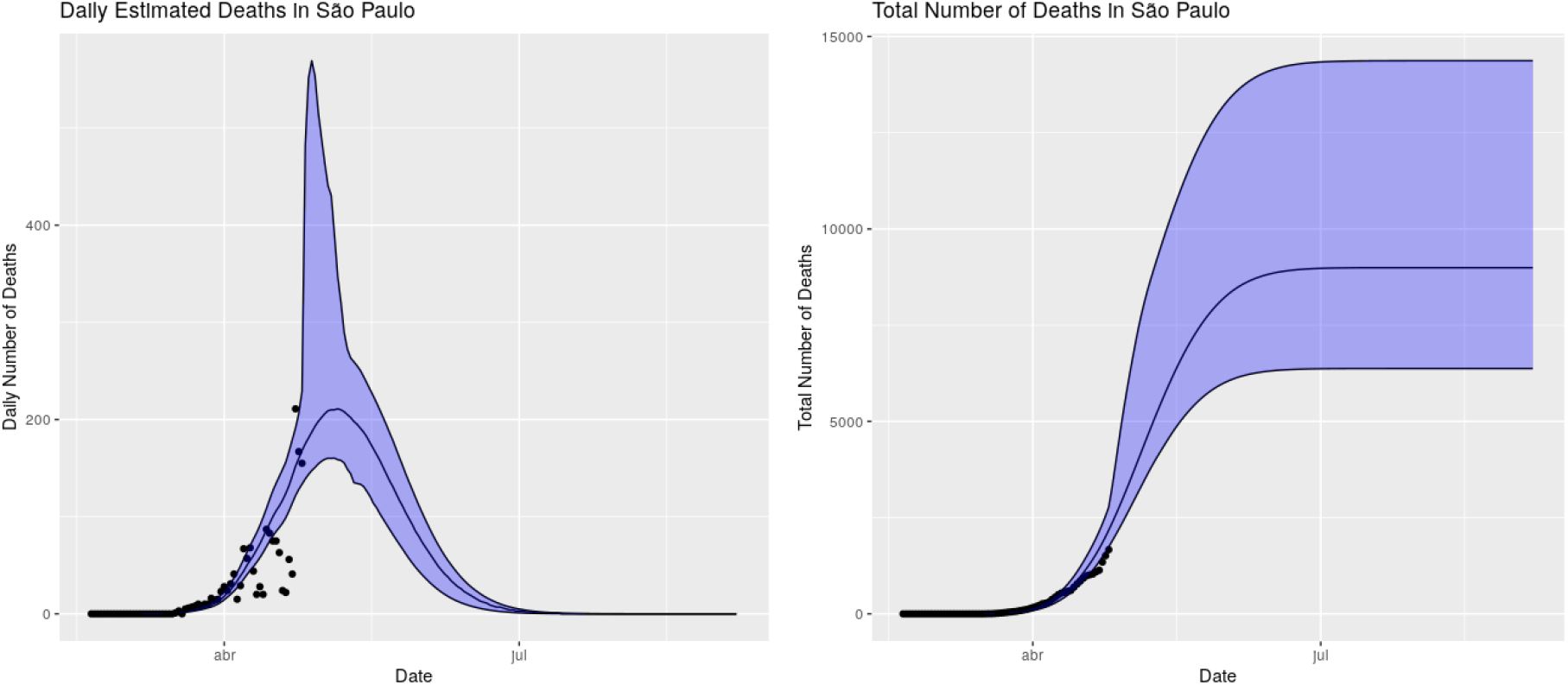
Fitted model and predicted values until the end of August.

One of our main goals with this model is to verify the usage of the ICU-beds to assess if the state will attain is maximum capacity, which would imply in lack of ICU beds for severe patients. To this end, we assumed that 30% of the ICU beds were occupied for other reasons, so 70% of the ICU beds were available to COVID-19 patients.

We see, in Figure 6 that the model suggests that São Paulo will not attain its maximum capacity of ICU beds if the current trend maintains to the long-term, such as the current mitigation and/or current hygiene and sanitary habits of São Paulo inhabitants.

**Figure 6:**
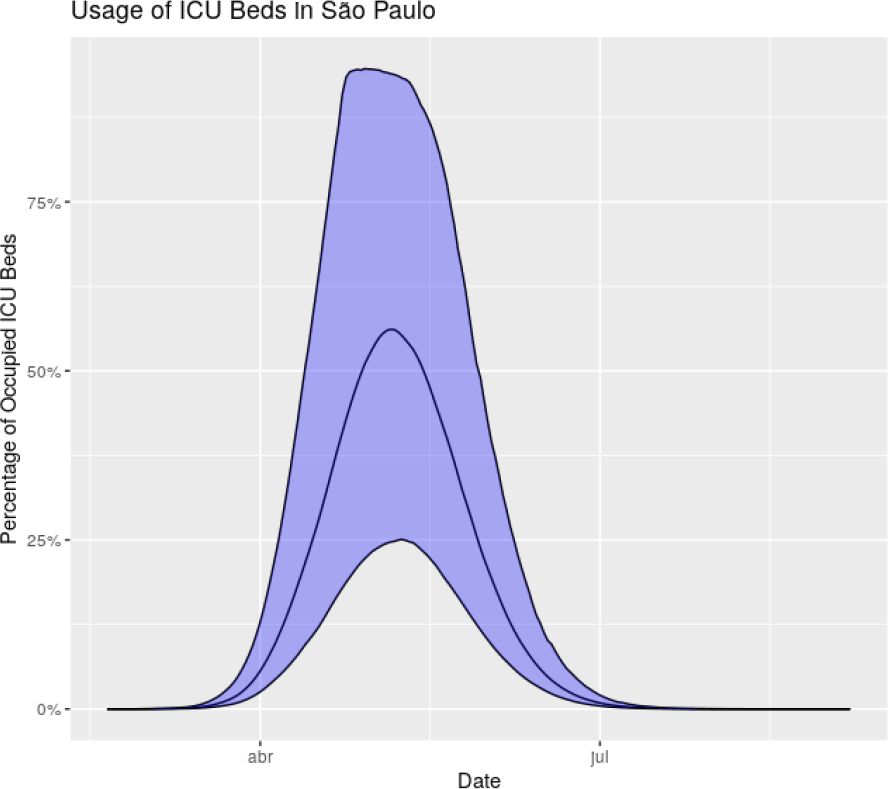
Estimated usage of ICU beds in São Paulo.

Finally, in Figure 7 we provide the estimated number of infected individuals in the state of São Paulo, Brazil. It is important to notice that whenever the number of infected individuals is away from the the total population, the dynamics of the model is the same for any choice of probability of need ICU care, so the number of infected individuals will vary by a large factor, but the shape and height of the curve for the estimated number of deceased individuals will remain the same even though the estimated number of infected individuals will vary a lot. That being said, our current choice of probabilities of need ICU-level care, as discussed in the previous sections, entails an estimated number of infected individuals of approximately 2.4 million individuals (median) in April 25, 2020, and reaching a total of 4 million individuals in the end of August, 2020 (median). These figures are much higher than the current estimated figures seen in other models, but as discussed in the introduction, we believe this number is closer to reality.

**Figure 7:**
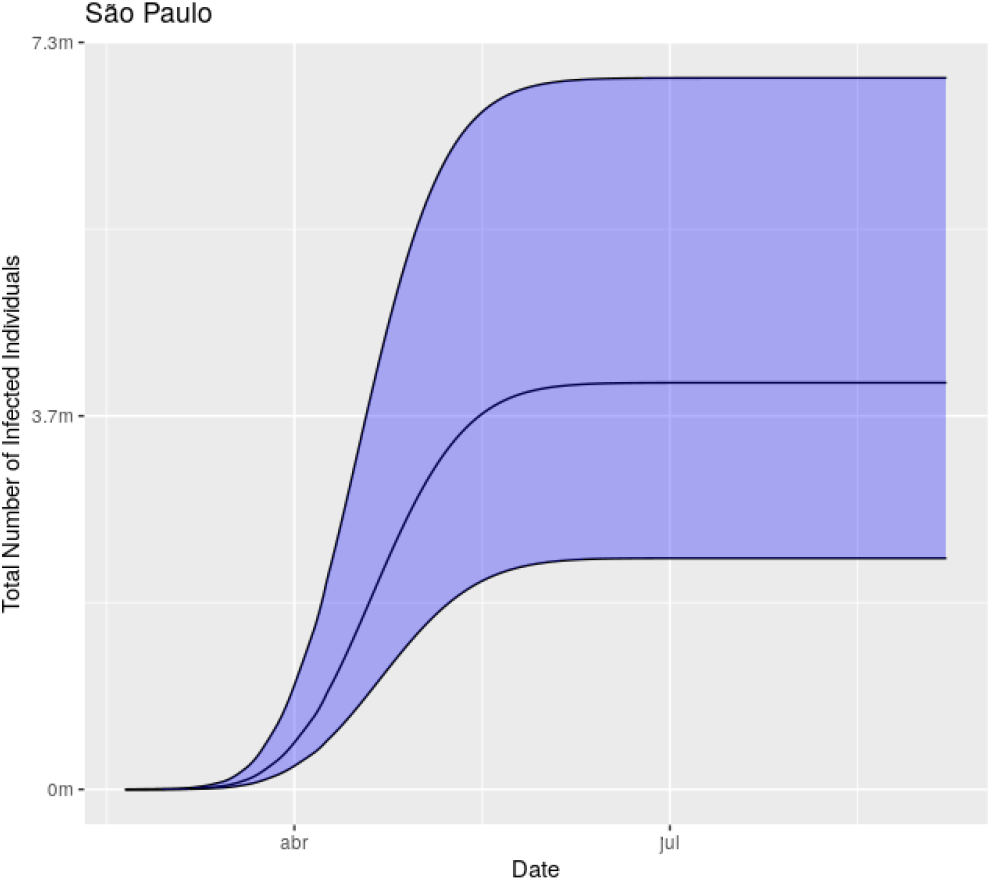
Estimated number of infected individuals by COVID-19 in São Paulo.

### 4.2 Model fitting in the state of São Paulo - separating public and private ICU beds

We will now provide results obtained when considering the previous model separating the usage of ICU beds for those who have health insurance plans from those who have not.

The proportion of people with health insurance plan was obtained in [40].

We considered the same calibration for (3) in this model to account for ICU occupation.

In Figures 8 and 9 we provide the predicted values for the next seven days and the long-term predictions, respectively. We observe that the results for the state of São Paulo are very similar to those without separation of the ICU beds. Also, notice that the “peaks” are expected to occur around the same days to those without separation. The median of the total number of deaths, with separation, is expected to be at around 9000 deaths, with 2.5% quantile being around 6600 deaths and the 97.5% quantile being around 13350 deaths.

**Figure 8:**
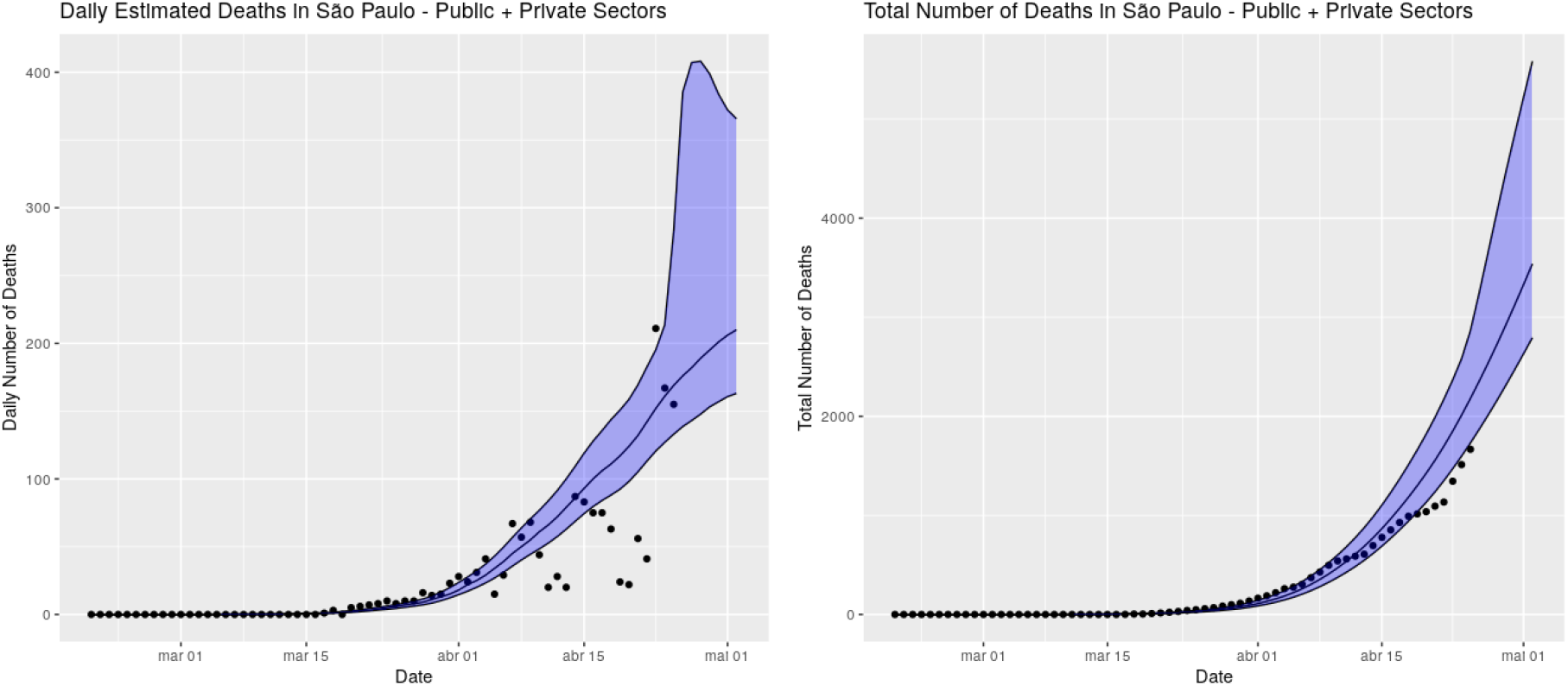
Fitted model and predicted values for the next seven days considering separation of ICU beds from the public and private sectors.

**Figure 9:**
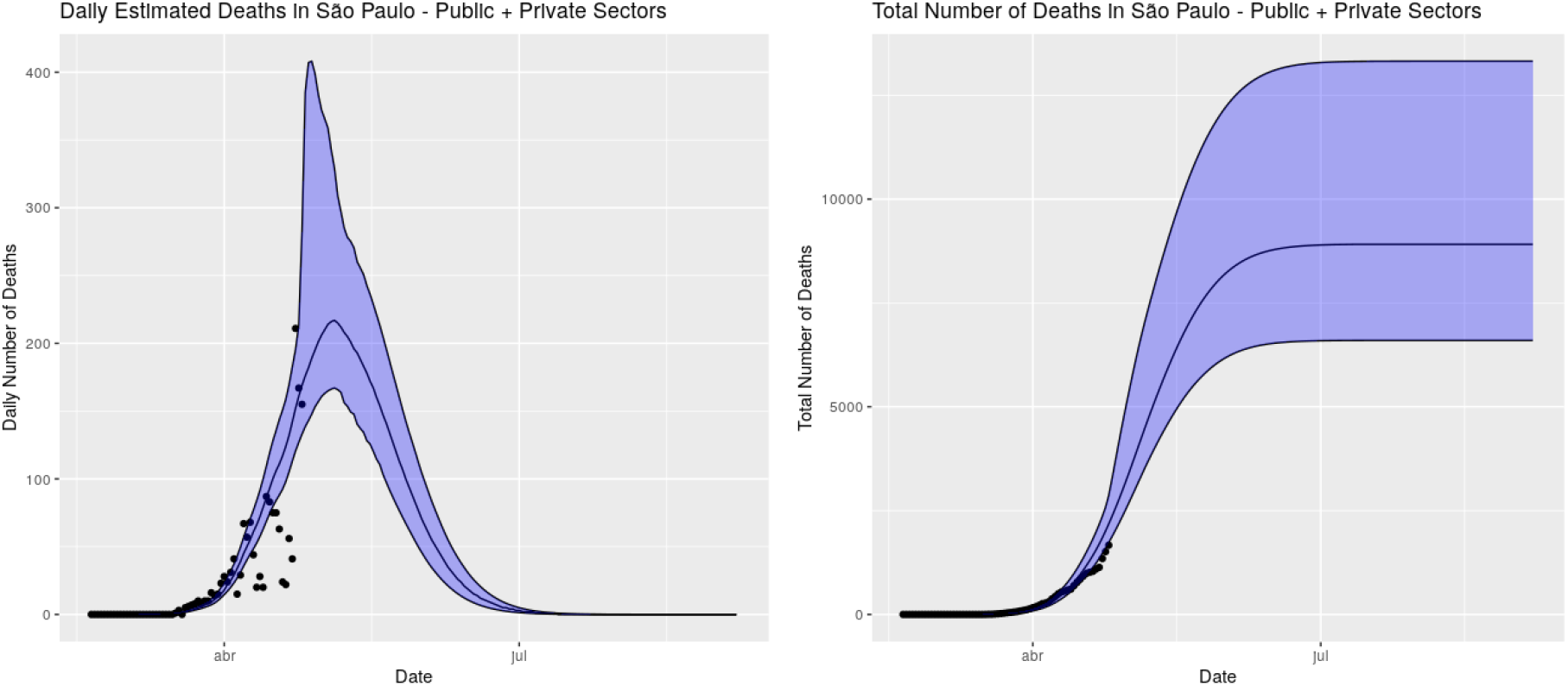
Fitted model and predicted values until the end of August considering separation of ICU beds from the public and private sectors.

It is noteworthy and not entirely intuitive that the higher band of the total deaths in the scenario separating the public and private ICU beds are lower than the higher band of the total number of deaths in the scenario without such separation. Our interpretation is that when the situation becomes very bad, in the sense that a lot of people are needing ICU-level care, if there is no separation, once the ICU beds are fully occupied, anyone who needs a bed will necessarily die. On the other hand, having separation of the beds, due to the high number of private ICU beds in São Paulo, it is likely that everyone who has health insurance plan will have an available bed, so only the portion of the population without health insurance plan will die in such a bad situation. However, this is a very improbable situation. In the most probable situations the separation increases the total number of deaths.

In Figure 10 we observe that the in the median scenario we expect the usage of the ICU beds in the public sector to be high, but we do not expect to have shortage of ICU beds in the public sector. The usage of ICU beds in the private sector are expected to be at a very comfortable level throughout the forecast period.

**Figure 10:**
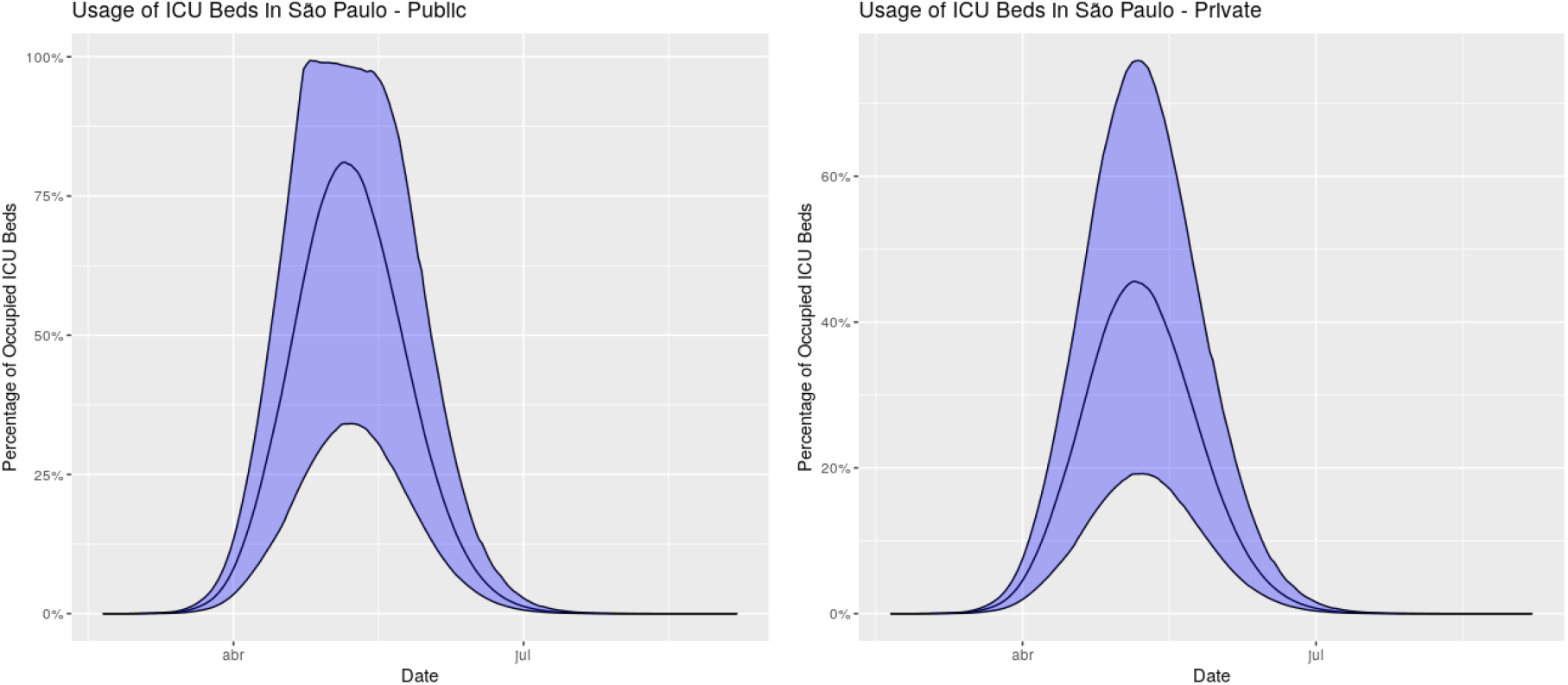
Estimated usage of ICU beds in São Paulo from the public and private sectors.

Finally, in Figure 11 we provide the estimated number of infected individuals in the state of São Paulo, Brazil in the case of separated ICU beds. Our current choice of probabilities of need ICU-level care, as discussed in the previous sections, entails an estimated number of infected individuals of approximately 2.5 million individuals (median) in April 25, 2020, and reaching a total of 4.15 million individuals in the end of August, 2020 (median). These figures are very close to the figures obtained without separation of beds.

**Figure 11:**
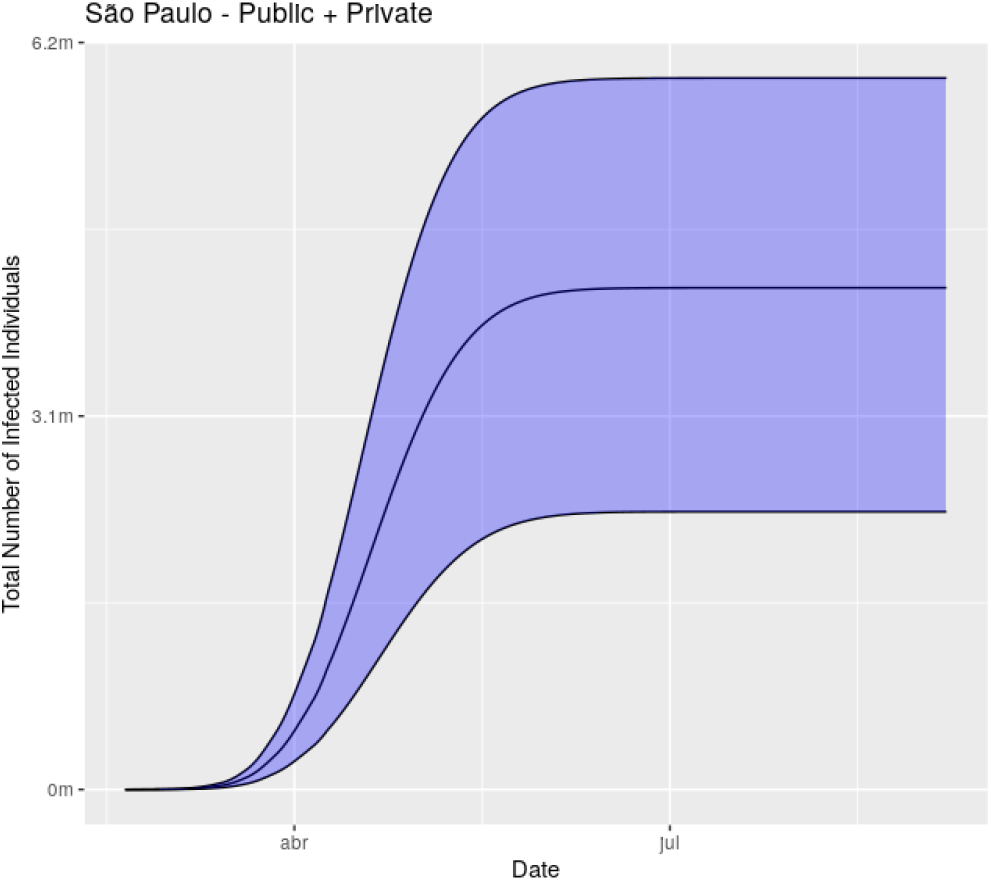
Estimated number of infected individuals by COVID-19 in São Paulo considering separation of ICU beds from the public and private sectors.

### 4.3 Model fitting in the regions of Brazil

We tried to avoid aggregating the data to fit the model. Instead, we tried to fit several models, one for each state in Brazil. Due to poor data, we were not able to fit all of them separately. So, instead, we fitted one model for each region of Brazil.

We will now describe the groups:

a. Northern region:
  - Amazonas, Roraima, Acre, Rondônia, Tocantins, Amapá, Pará
b. Northeastern region:
  - Ceará, Pernambuco, Rio Grande do Norte, Alagoas, Sergipe, Piauí, Paraíba, Bahia
c. Southeastern region:
  - São Paulo, Rio de Janeiro, Espírito Santo, Minas Gerais
d. Central-Western region:
  - Mato Grosso do Sul, Mato Grosso, Distrito Federal, Goiás
e. Southern region:
  - Paraná, Santa Catarina, Rio Grande do Sul

For all the regions, the model converged. The models also had good performance in a time-series cross-validation with respect to three days ahead forecasting.

In Figures 12 and 13 we have, respectively, the daily and total estimated deaths for all regions of Brazil. It is noteworthy that the daily number of reported deaths is very noisy, but the total number of deaths is smoother and the model provides an overall good fit. The model also compensate the assumed under-reported deaths across the different regions from Brazil (this is controlled through the choice of the sign in equation (3)). In the southeastern region we used the same calibration of (3) used for São Paulo. Notice that we have low data from the northern, southern and central-western regions, so we expect our predictions to improve a lot once new and better data are available. Finally, recall that in this situation we are considering that there is no separation between ICU beds from the public and private sectors.

**Figure 12:**
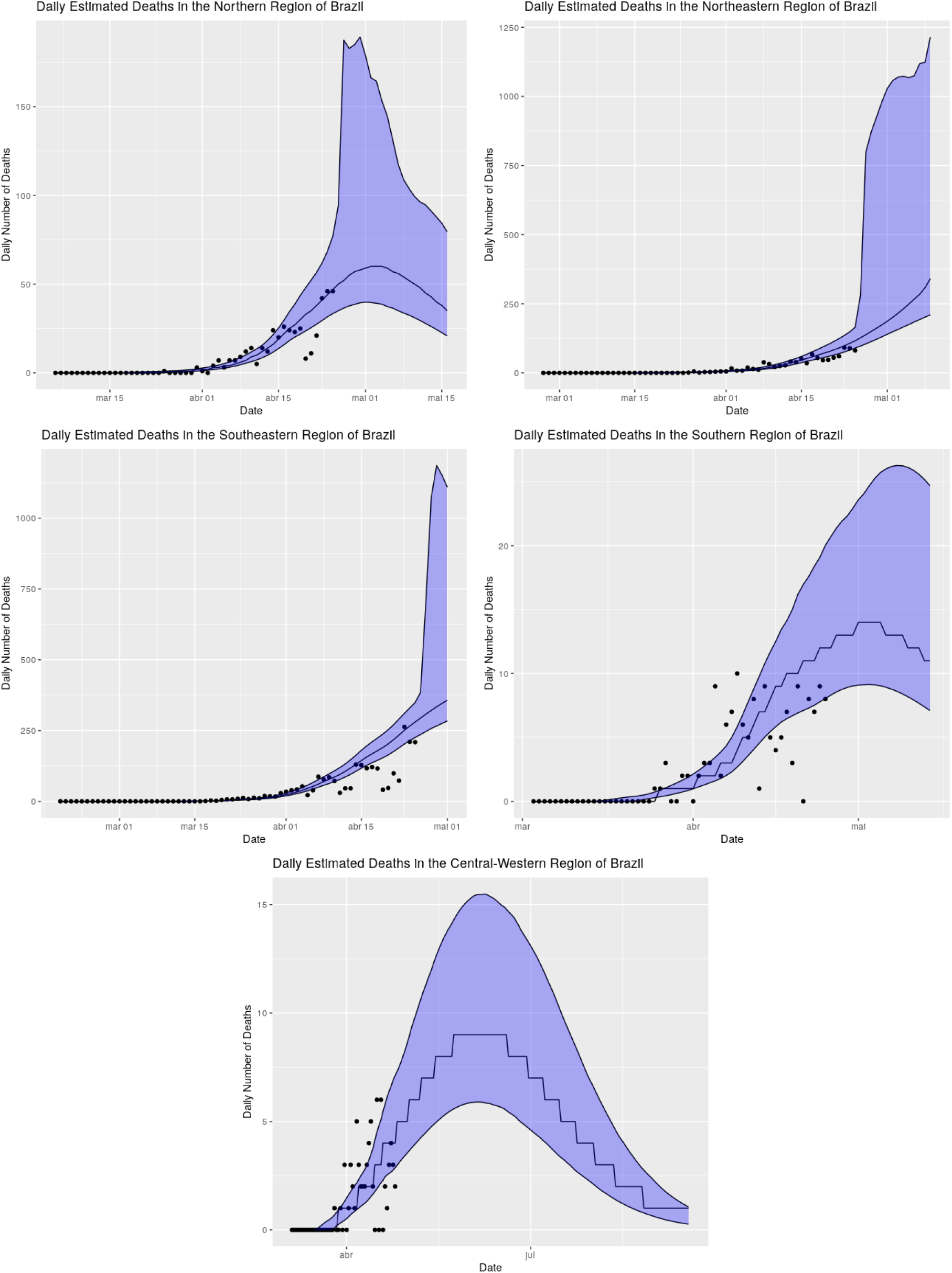
Fitted model and predicted values for the next seven days across the different regions from Brazil.

**Figure 13:**
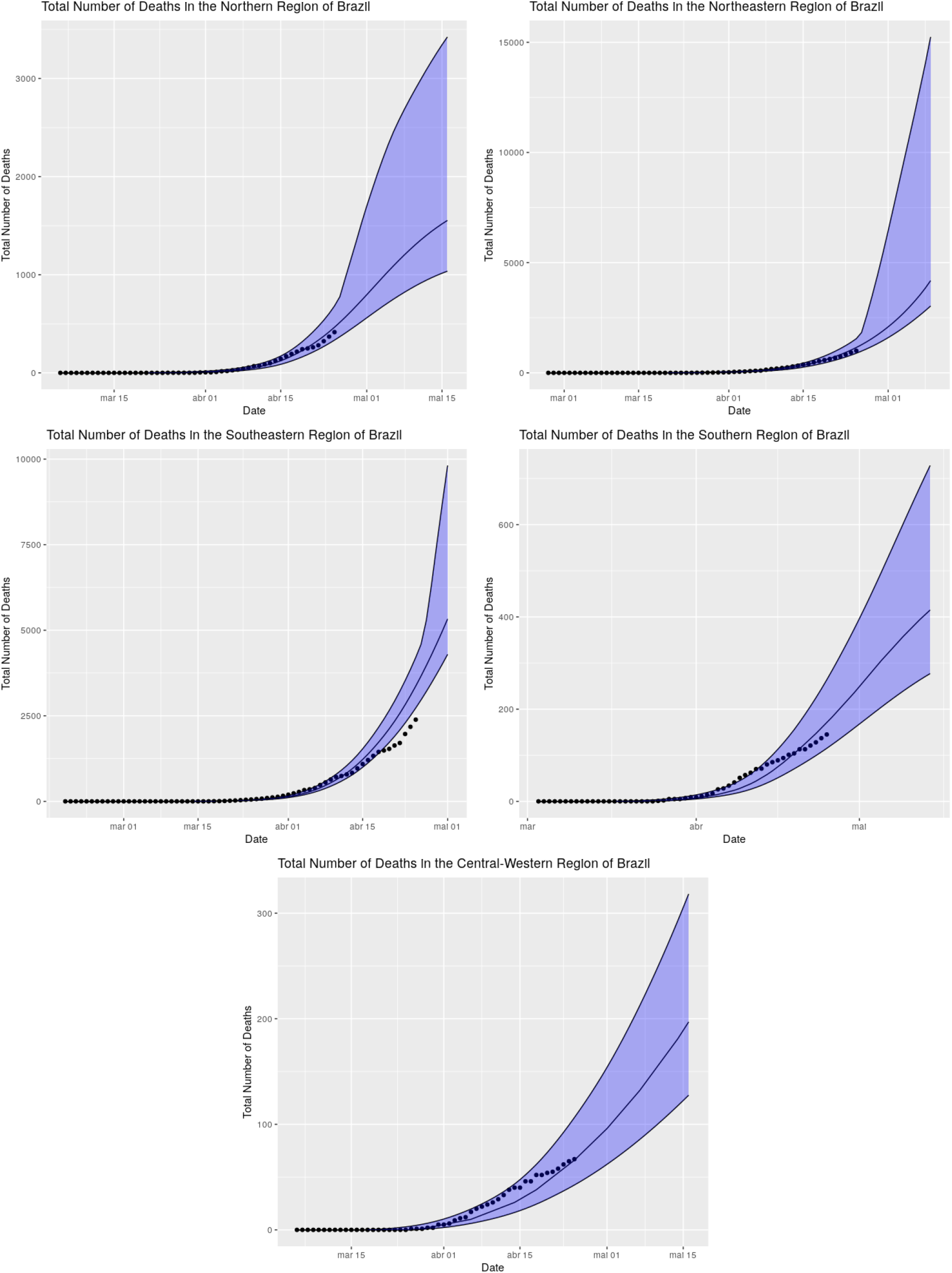
Fitted model and predicted values for the next seven days across the different regions from Brazil.

In Figures 14 and 15 we have the daily and total estimated deaths for all regions of Brazil up to the end of August, 2020. The models suggest that the “peak” of the northern region will occur around from April 26th to May 10th, 2020. For the northeastern region, the “peak” is estimated to occur from 11th to 22th of May, 2020. For the southeastern region the “peak” is expected to occur from May 3rd to May 11th. For the southern region the “peak” is expect to occur from 1st to 5th of May, 2020, whereas for the central-western region the “peak” is expect to occur from June 1st to June 19th. With respect to the number of deaths, the median of the total number of deaths in the northern region is around 1900. For the northeastern region, the median of the total number of deaths is around 19000 (as we will see below, the model expects this number to be very high due to lack of available ICU beds). For the southeastern region, the median of the total number of deaths is around 16700. Finally, for the southern the median of the total number of deaths is around 600, whereas for the central-western region the median of the total number of deaths is around 850. We would like to recall that all these estimates assume that the current policies and the behavior of the population are maintained throughout the forecast period.

**Figure 14:**
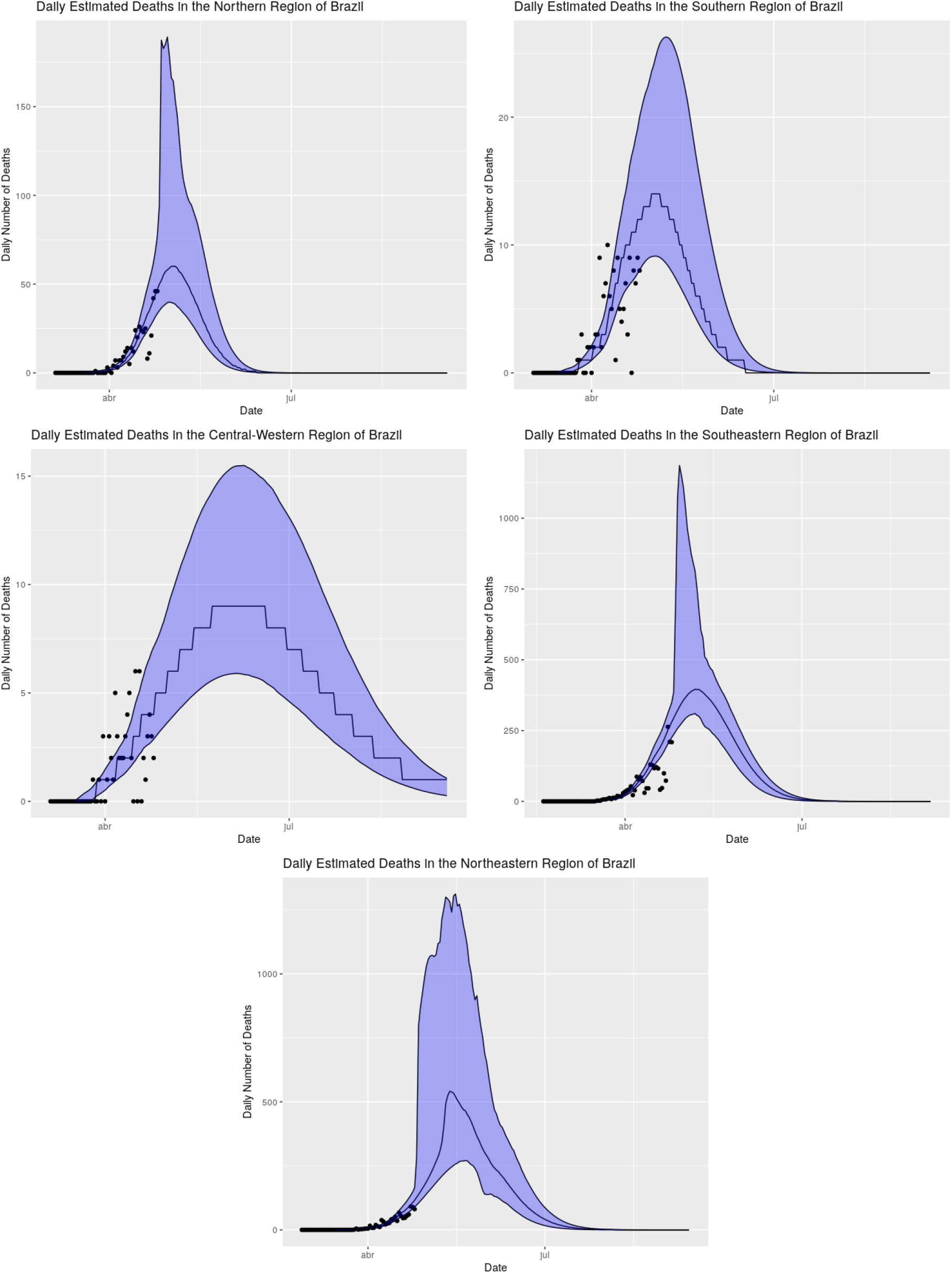
Fitted model and predicted values for the next seven days across the different regions from Brazil.

**Figure 15:**
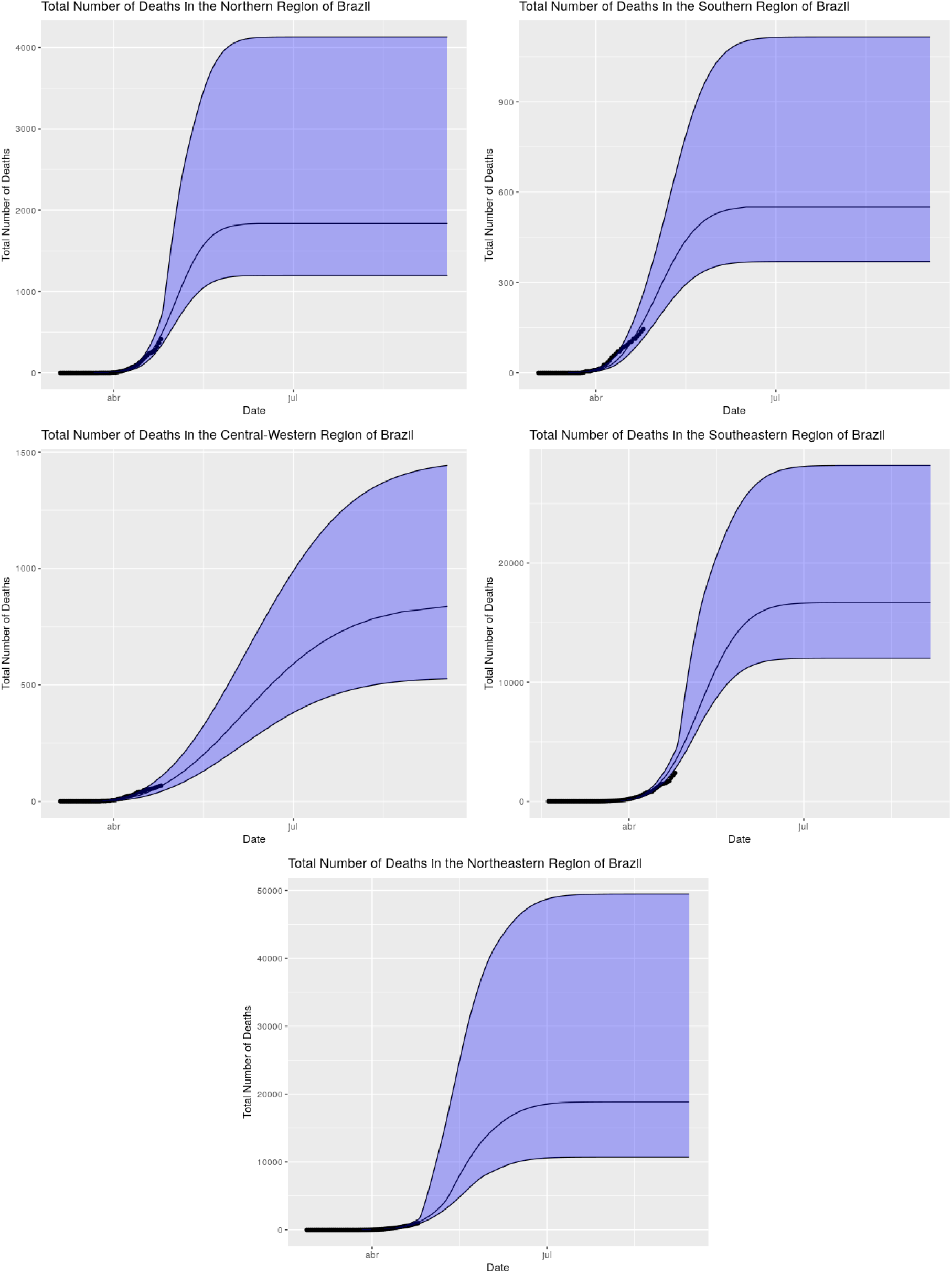
Fitted model and predicted values until the end of August across the different regions from Brazil.

In Figure 16 we see the percentage of usage of ICU beds across the different regions of Brazil. We see clearly that the northern and northeastern regions of Brazil will suffer from lack of available ICU beds, whereas the southeastern, southern and central-western regions appear to be able to handle. The central-western region has the federal district, which is an outlier, having a large number of ICU beds. These regions, south and central-west, deserve further attention (which will be done in the future), since they do not have enough data for a thorough study.

**Figure 16:**
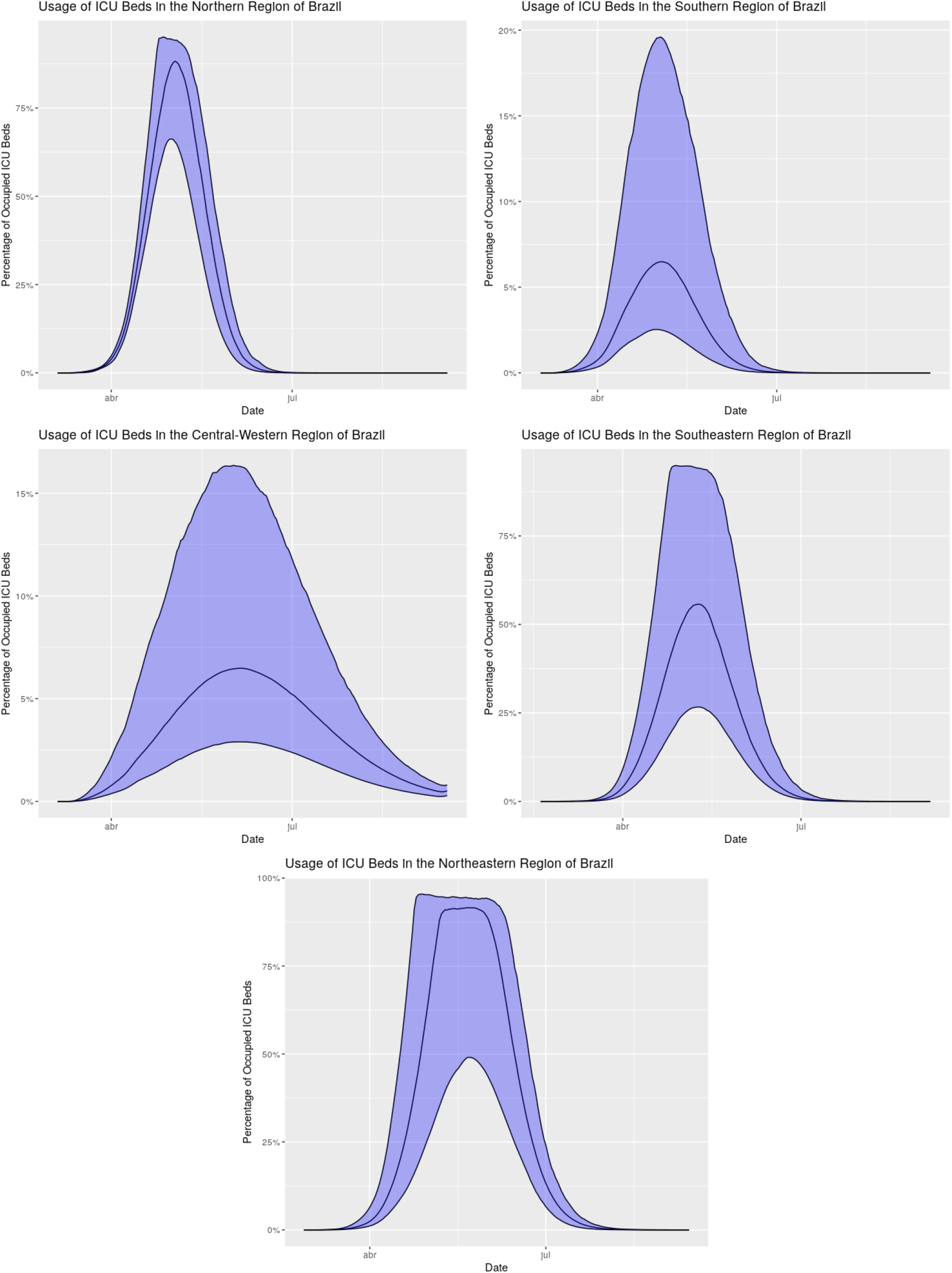
Estimated usage of ICU beds across the different regions of Brazil.

Finally, in Figure 17 we provide the estimated number of infected individuals across the regions of Brazil. Our model indicates that if the current policies and population behavior are maintained until the end of August, the proportion of immunized individuals in the northern region will be 5.3%; in the northeastern region will be around 13.7%; in the southeastern region will be around 8.8%; in the southern region will be around 1% finally, the model expects the proportion of immunized individuals in the central-western region at the end of august to be around 2.4%.

**Figure 17:**
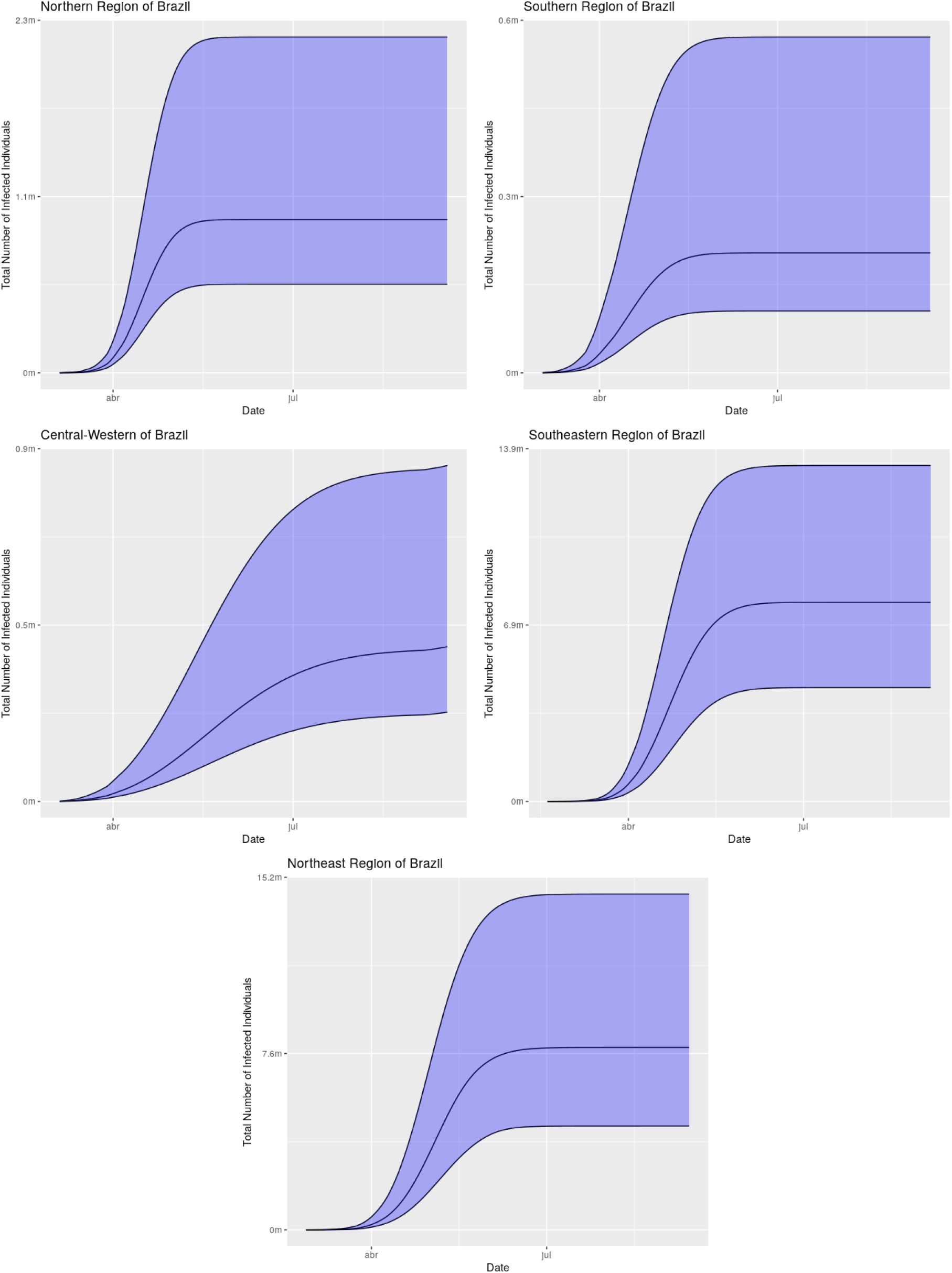
Estimated number of infected individuals across the different regions of Brazil.

### 4.4 Model fitting in the regions of Brazil - separating public and private ICU beds

We will now provide results obtained when considering the regions of Brazil separating the usage of ICU beds for those who have health insurance plans from those who have not.

The proportion of people with health insurance plan was obtained in [40].

In the southeastern region, we considered the same calibration for (3) in this model to account for ICU occupation.

In Figures 18 and 19 we provide the predicted values for the next seven days of the daily deaths and total number of deaths, respectively. We observe that the results for the different regions are similar to those without separation of the ICU beds considering the predictions for the next 7 days.

**Figure 18:**
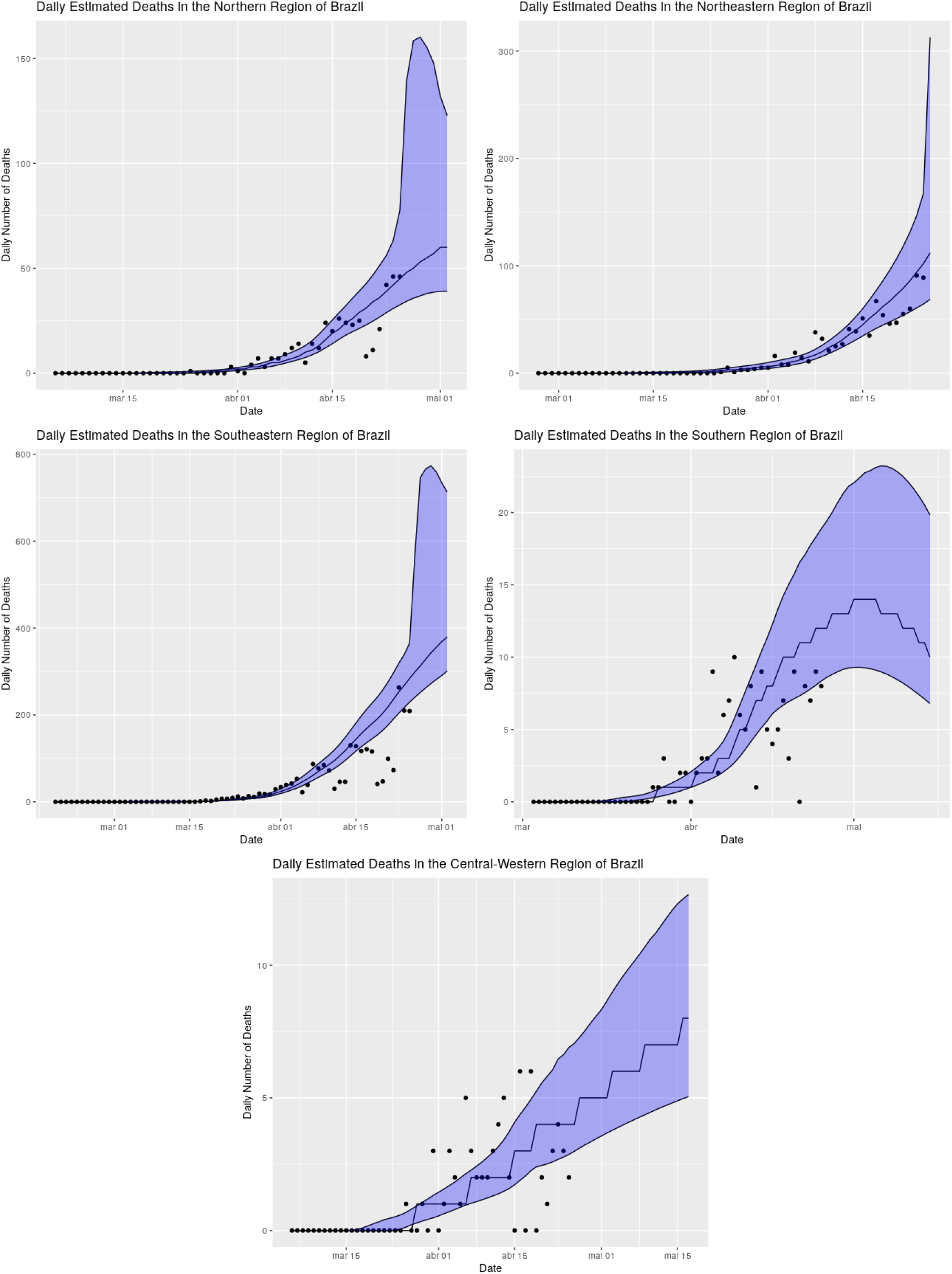
Fitted model and predicted values for the next seven days across the different regions from Brazil considering separation of ICU beds from the public and private sectors.

**Figure 19:**
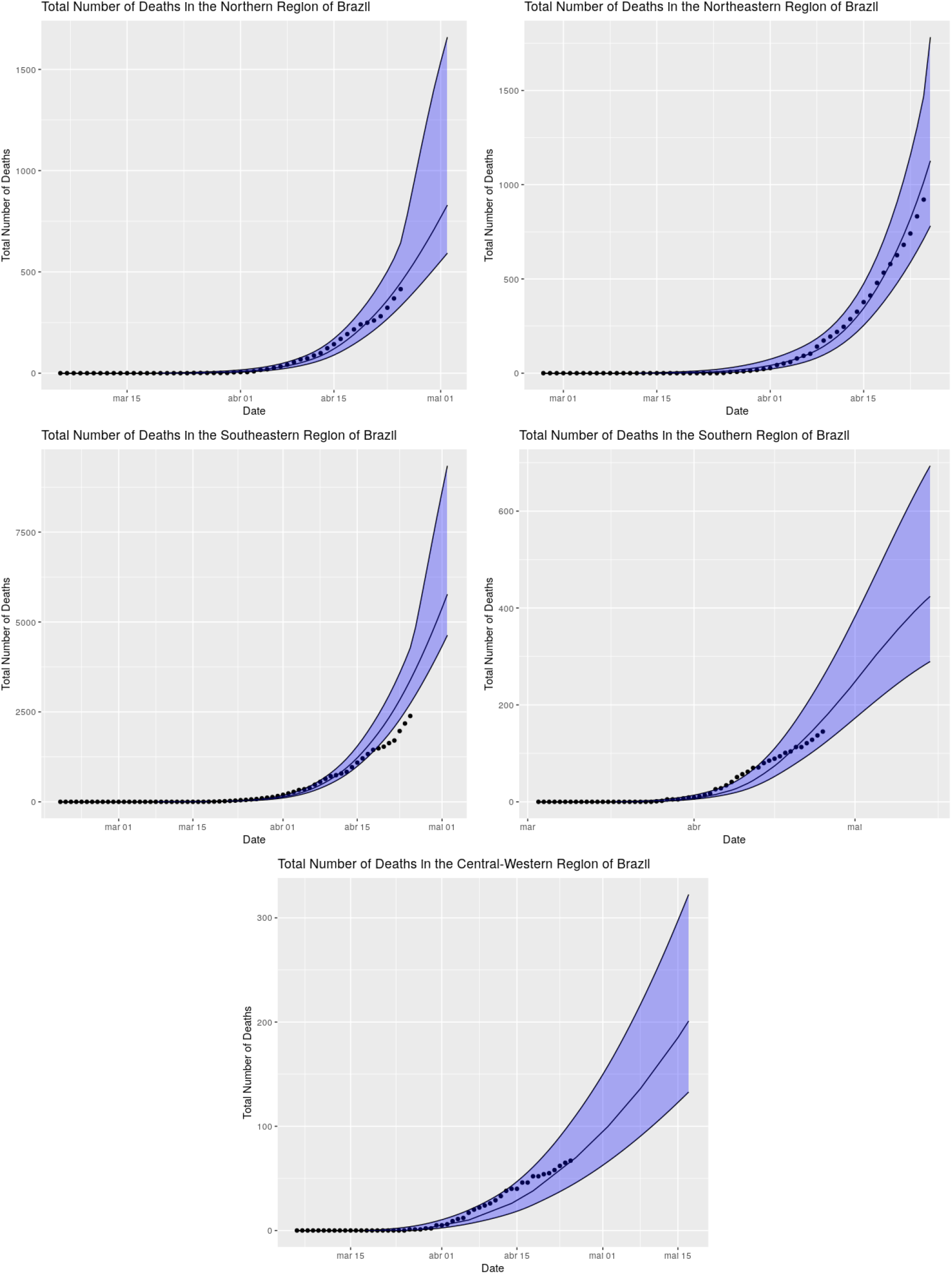
Fitted model and predicted values for the next seven days across the different regions from Brazil considering separation of ICU beds from the public and private sectors.

We now move to long-term predictions. To this end, we provide in Figures 20 and 21 the daily and total estimated deaths for all regions of Brazil up to the end of August, 2020. Notice that the “peaks” are expected to occur around the same days to those without separation. For the northern, southern and central-western regions, the total number of deaths considering the separation is very close to the scenario where there is no separation. For the northern region, the reason is the low number of ICU beds, whereas for the southern and central-western regions, the reason is the high number of ICU beds. For the southeastern region, the total number (median) of deaths is around 17650, whereas for the northeastern region the total number (median) of deaths is around 21800.

**Figure 20:**
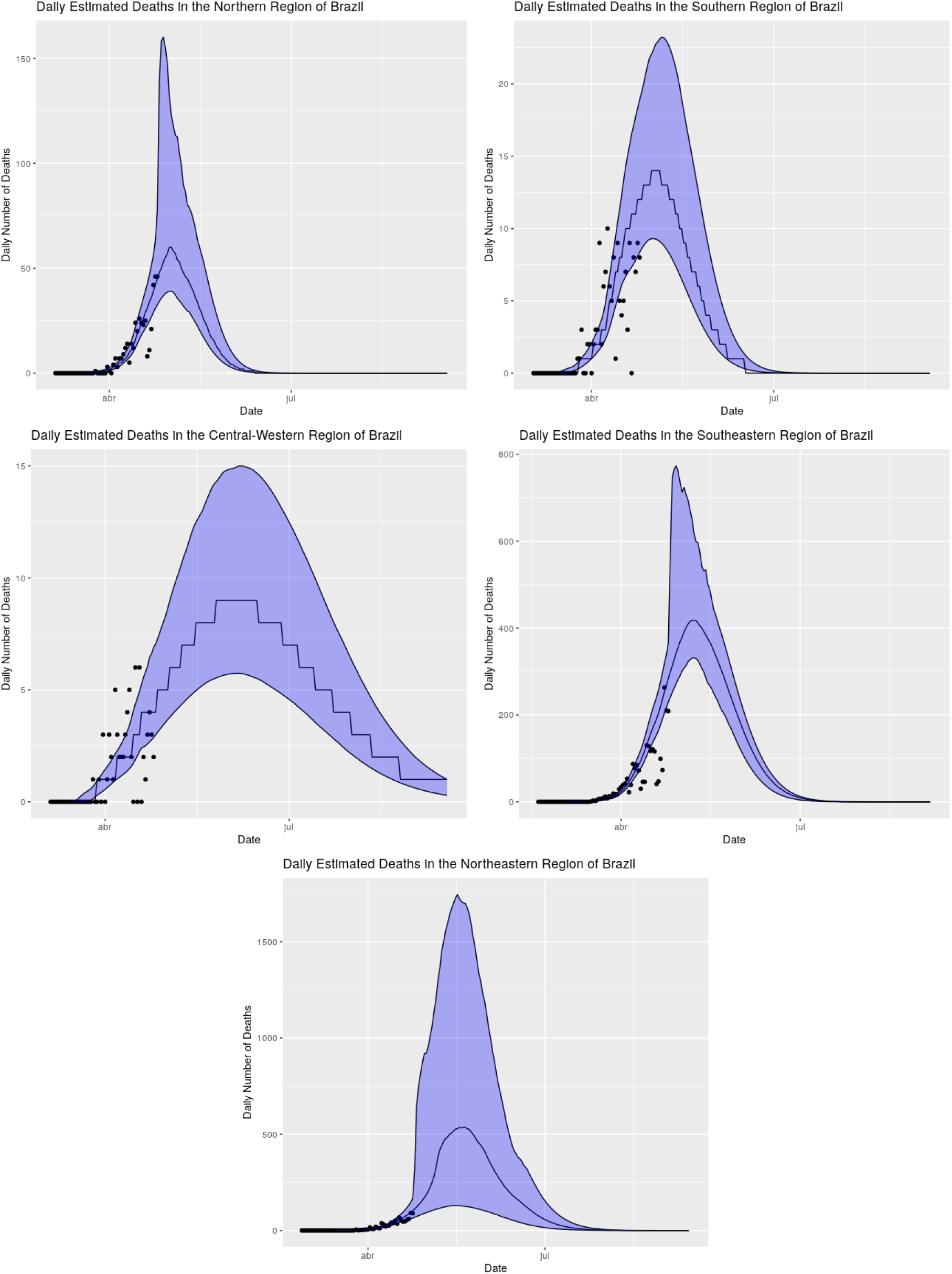
Fitted model and predicted values for the next seven days across the different regions from Brazil considering separation of ICU beds from the public and private sectors.

**Figure 21:**
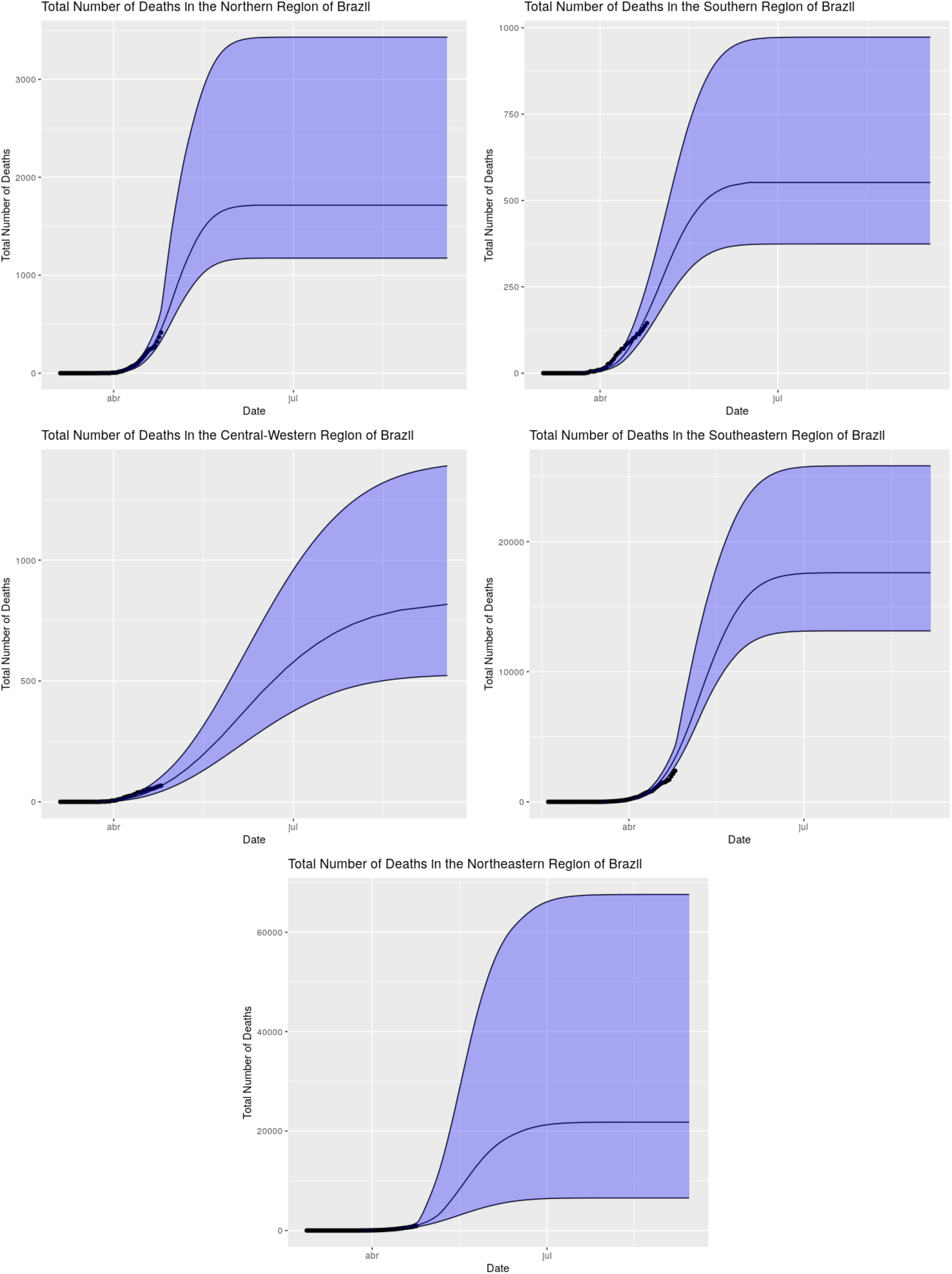
Fitted model and predicted values until the end of August across the different regions from Brazil considering separation of ICU beds from the public and private sectors.

In Figures 22 and 23 we provide the usage of ICU beds from the public and private sectors, respectively. Observe that the in the scenario of separation of ICU beds, the model indicates that the northern, south-eastern and northeastern regions may not have enough public ICU beds for those who need it. The southern and central-western regions seem to be in a comfortable situation with respect to ICU beds. All the regions seem to have enough ICU beds for those who have health insurance plans if the separation of ICU beds hold.

**Figure 22:**
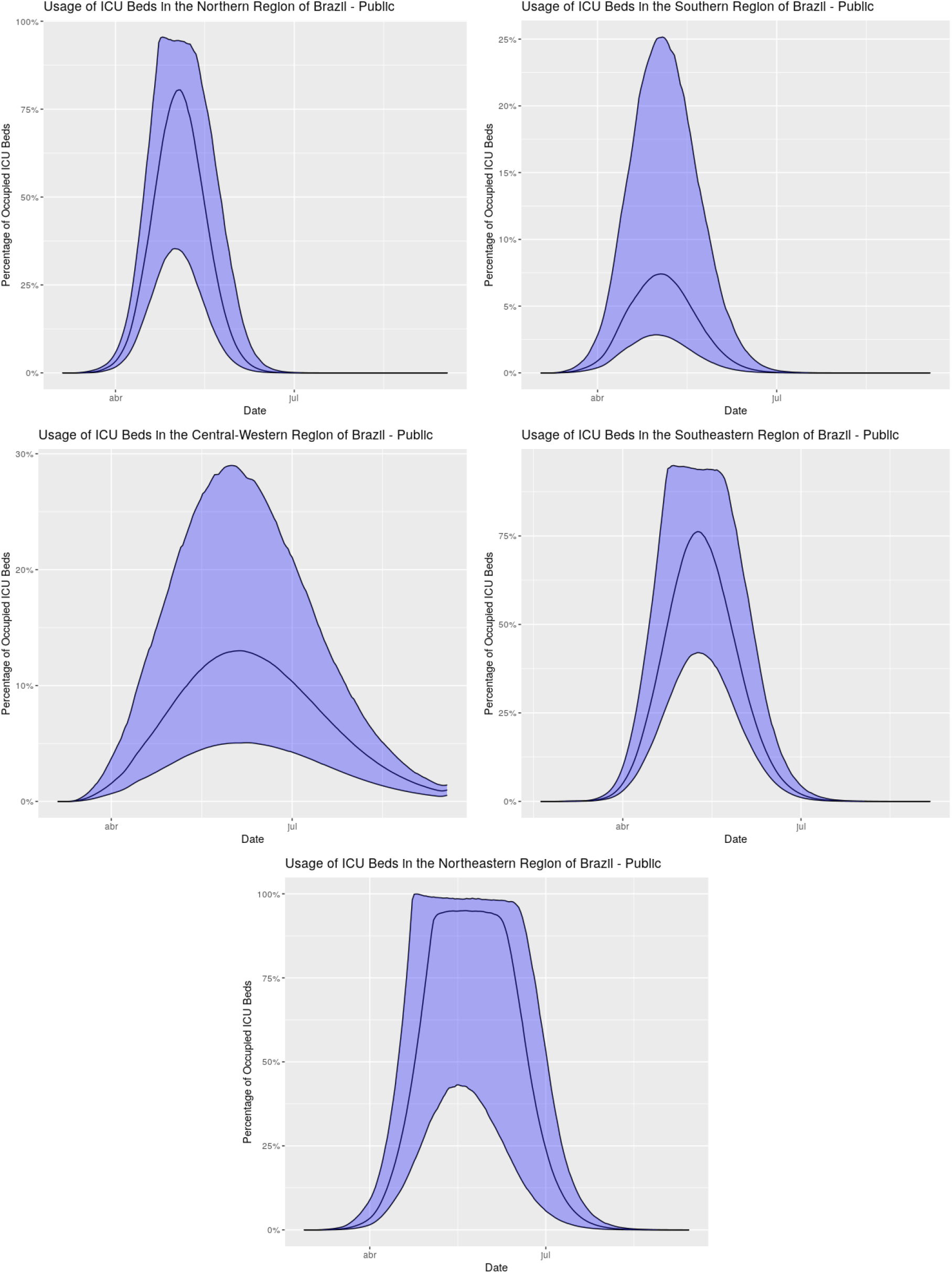
Estimated usage of ICU beds from the public sector across the different regions of Brazil.

**Figure 23:**
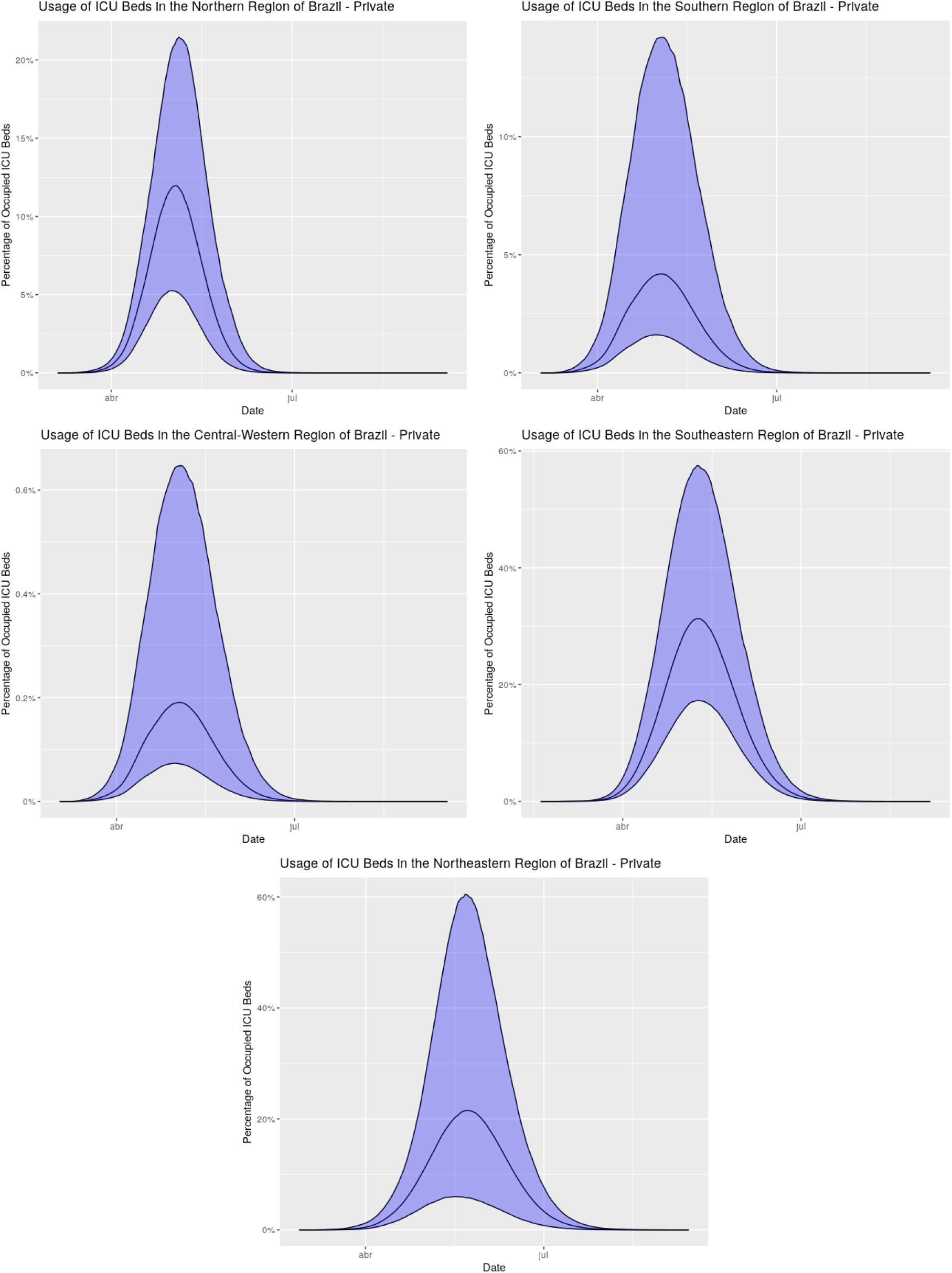
Estimated usage of ICU beds from the private sector across the different regions of Brazil.

**Figure 24:**
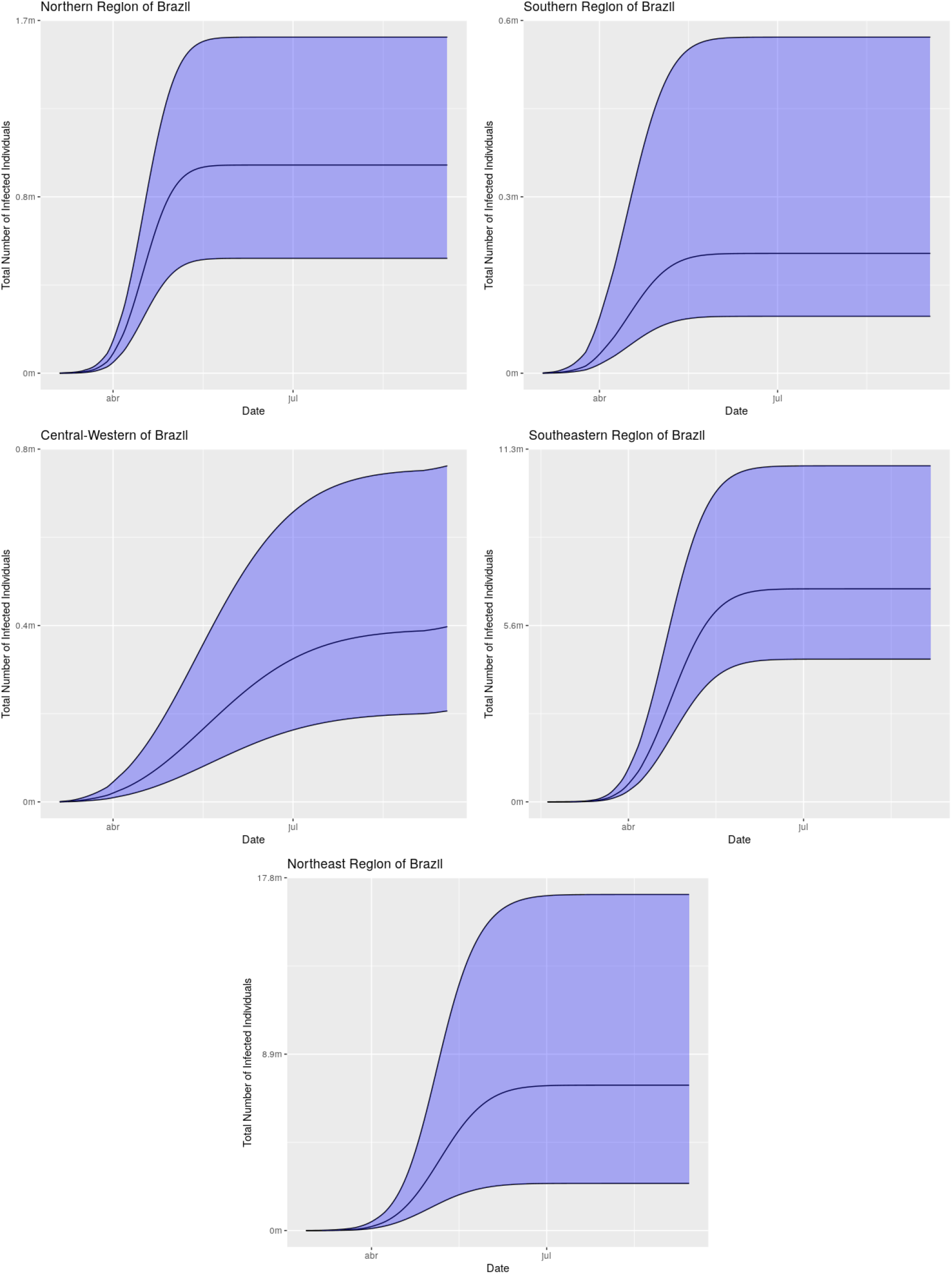
Estimated number of infected individuals across the different regions of Brazil considering separation of ICU beds from the public and private sectors.

Finally, in Figure 11 we provide the estimated number of infected individuals for the different regions of Brazil in the case of separated ICU beds. The estimated numbers are very close to those obtained without separation of beds.

### 4.5 Model fitting in Brazil

We obtained predictions to Brazil by summing the corresponding figures throughout the different regions of Brazil. We expect this result to be more reliable than to directly fit the aggregated data from Brazil due to its heterogeneity. The different regions from Brazil have very different age pyramids, ICU beds per 100 thousand inhabitants, percentage of population with access to private hospitals, etc. So we tried to break Brazil into approximately homogeneous parts (the regions). Actually, even the regions are heterogeneous, but unfortunately we do not have enough data to obtain a reasonably nice fit for each state.

In Figure 25 we present the fitted model and the predicted values for the number of deaths in Brazil for the next seven days.

**Figure 25:**
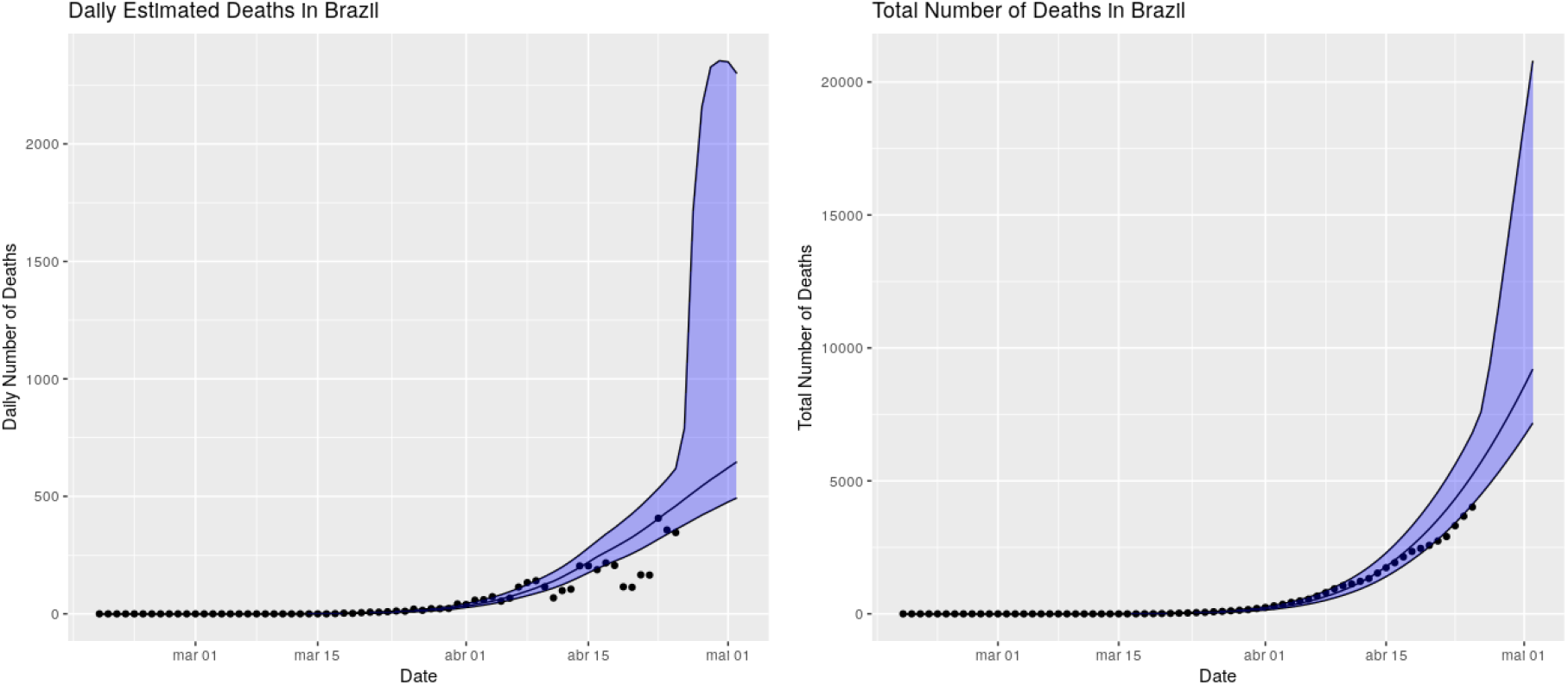
Fitted model and predicted values for the number of deaths in Brazil for the next seven days.

We will now provide our long-term predictions. In Figure 26 we provide the predictions of the model until the end of August. If the current policies and population behavior are maintained the “peak” of daily number of deaths is expected to occur from May 9th to May 20th, 2020. The “peak” is expected to be very long due to the fact that the “peaks” of the different regions are attained at different moments of time. The total number of deaths is being estimated at around 38800 (median) with 2.5% quantile being around 24900 deaths and the 97.5% quantile being around 84,500 deaths. These estimated totals vary a lot through the days due to the exponential nature of the model and the bad quality of the data seen in Brazil, but the model has been very consistent with the estimated date of the “peak”.

**Figure 26:**
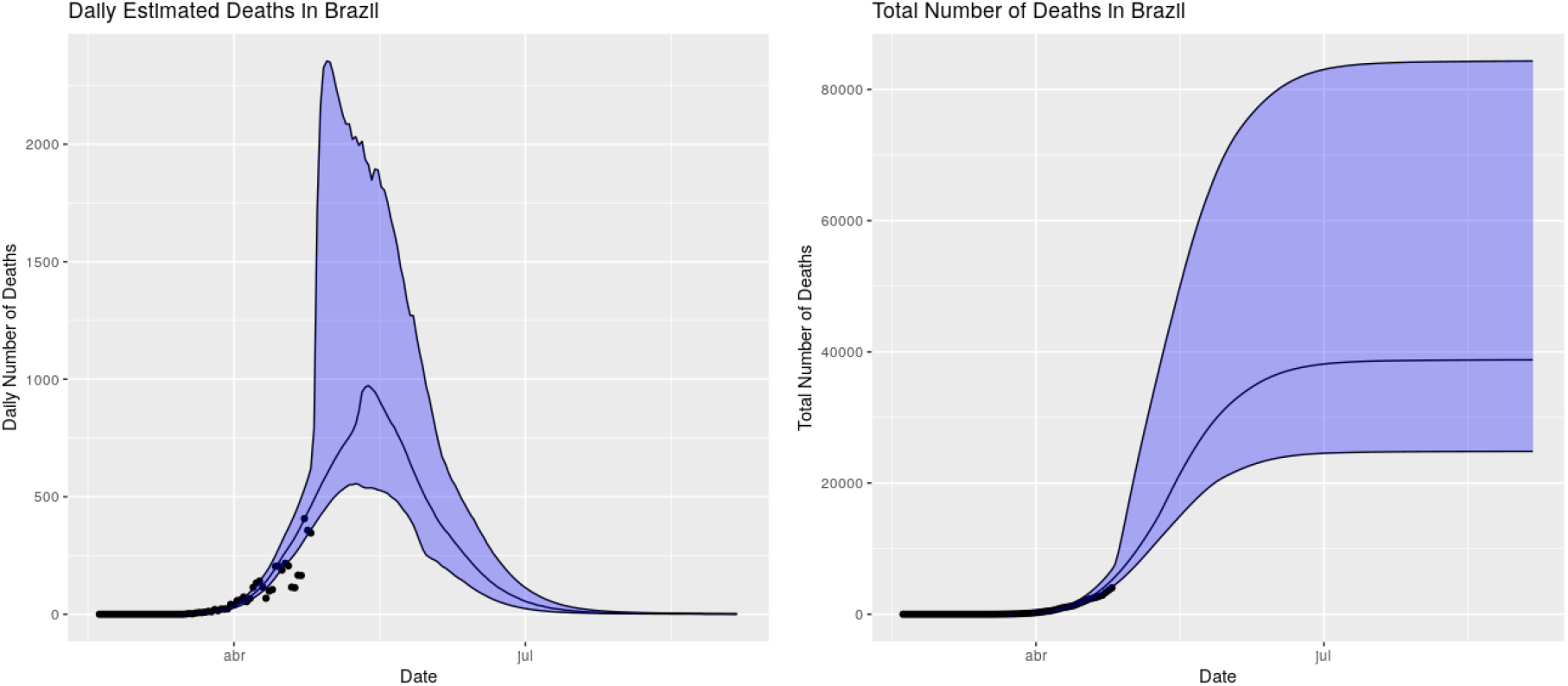
Fitted model and predicted values for the number of deaths in Brazil until the end of August.

We will not provide the usage of ICU beds for this case since there is no mobility between ICU beds.

Finally, in Figure 27 we provide the estimated number of infected individuals in Brazil. The model expect that if the current policies and population behavior are maintained throught the forecasted period, at the end of August, 2020, Brazil will have around 8.2% of its population immunized of COVID-19.

**Figure 27:**
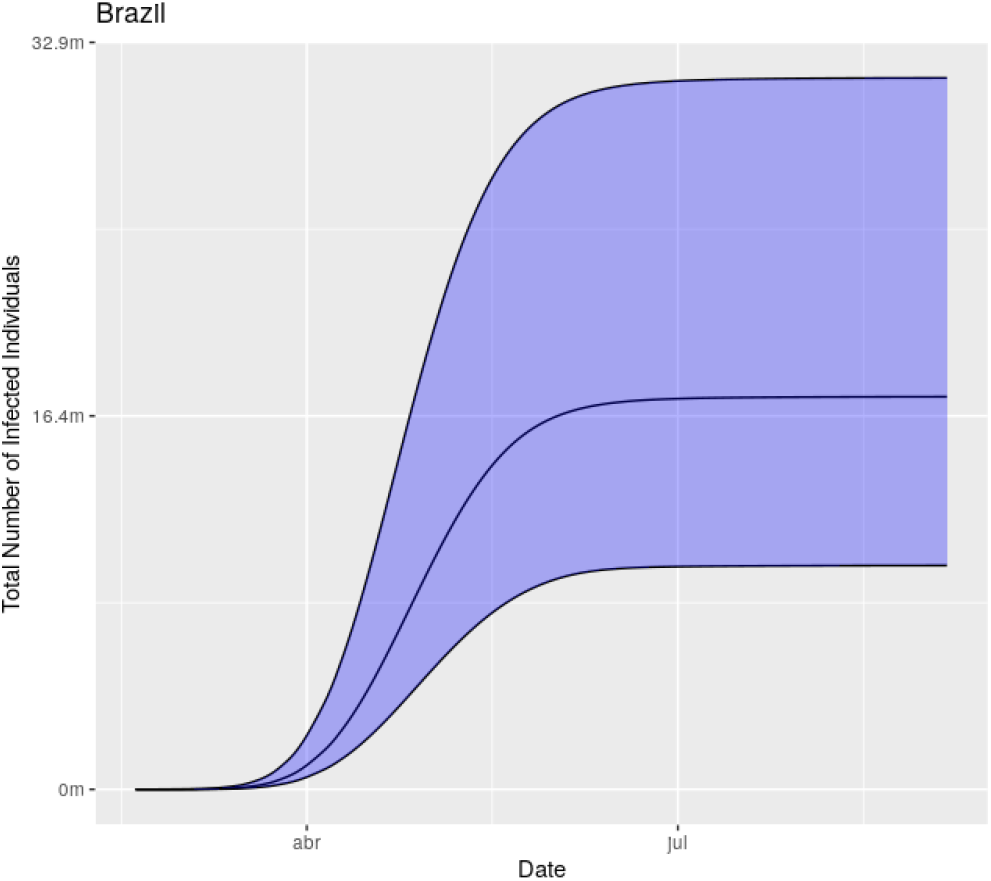
Estimated number of infected individuals by COVID-19 in Brazil.

### 4.6 Model fitting in Brazil - separating public and private ICU beds

We will now provide results obtained by summing the corresponding results for the different regions of Brazil when separating the usage of ICU beds for those who have health insurance plans from those who have not.

The proportion of people with health insurance plan was obtained in [40].

In Figure 28 we present the fitted model and the predicted values for the number of deaths in Brazil for the next seven days.

**Figure 28:**
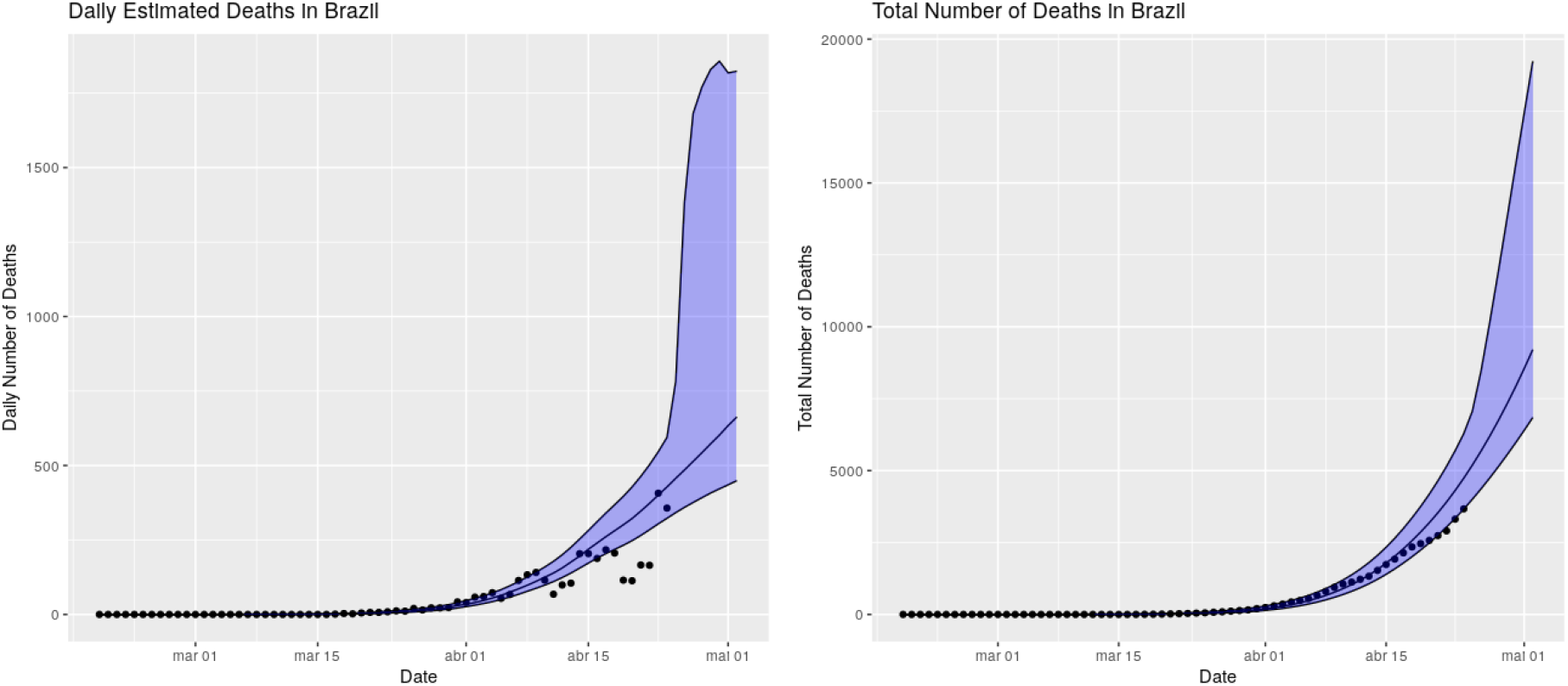
Fitted model and predicted values for the number of deaths in Brazil for the next seven days considering separation of ICU beds from the public and private sectors.

We will now provide our long-term predictions. In Figure 29 we provide the predictions of the model until the end of August. If the current policies and population behavior are maintained the “peak” of daily number of deaths is expected to occur from May 8th to May 19th, 2020. The “peak” is expected to be very long due to the fact that the “peaks” of the different regions are attained at different moments of time. The total number of deaths is being estimated at around 42500 (median) with 2.5% quantile being around 21800 deaths and the 97.5% quantile being around 99,200 deaths. These estimated totals vary a lot through the days due to the exponential nature of the model and the bad quality of the data seen in Brazil, but the model has been very consistent with the estimated date of the “peak”.

**Figure 29:**
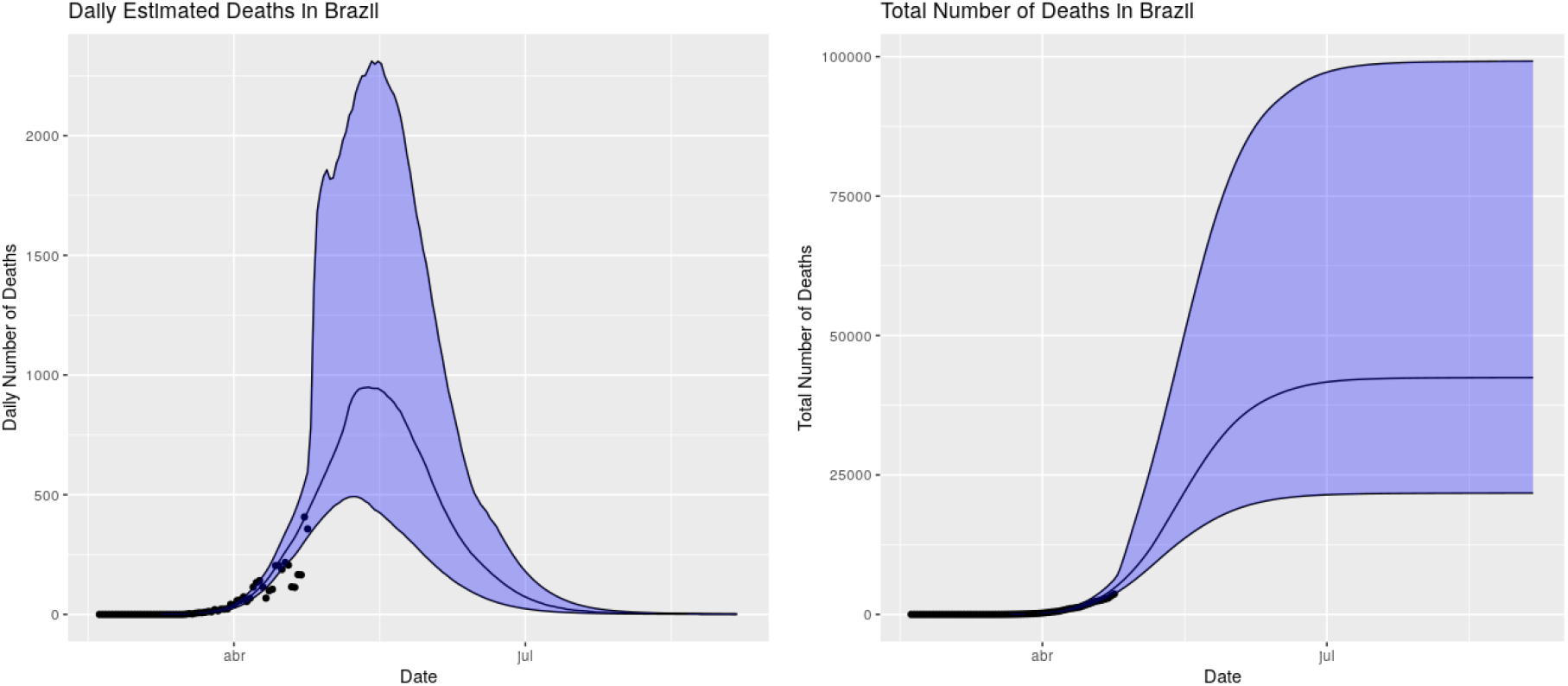
Fitted model and predicted values for the number of deaths in Brazil until the end of August considering separation of ICU beds from the public and private sectors.

We will not provide the usage of ICU beds for this case since there is no mobility between ICU beds.

Finally, in Figure 30 we provide the estimated number of infected individuals in Brazil. The model expect that if the current policies and population behavior are maintained throught the forecasted period, at the end of August, 2020, Brazil will have around 7.6% of its population immunized of COVID-19.

**Figure 30:**
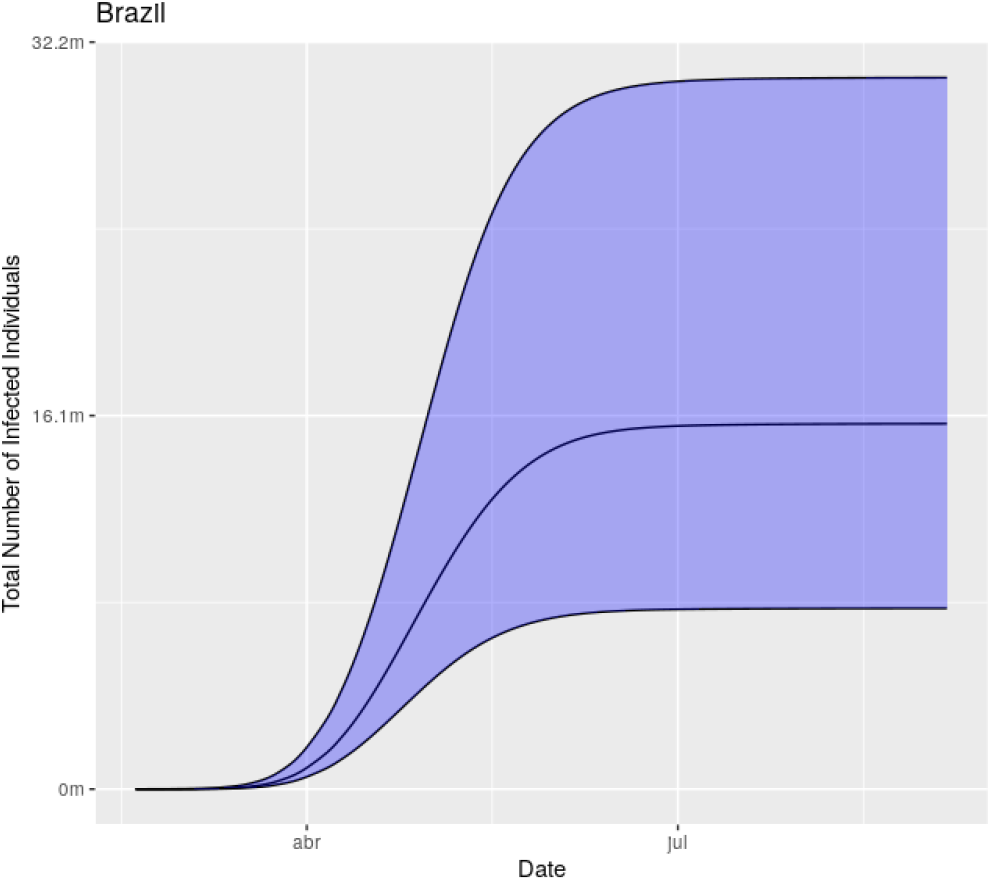
Estimated number of infected individuals by COVID-19 in Brazil considering separation of CU beds from the public and private sectors.

## 5 Discussion and Conclusions

Other mathematical models have been developed to predict the evolution of COVID-19 [41, 42, 34, 43]. As COVID-19 is a new disease, there is a great degree of uncertainty regarding its epidemiological variables [44]. The study by Ferguson et al. [34] assumed that 50% of the infected individuals could be asymptomatic. Recent evidence shows the possibility of a greater number of asymptomatic patients [44, 45, 26, 11]. Thus, we created a correction factor to the table described by Verity et al. [31] based on the work of Ioannidis et al. [22] and adapted it for the relative risk of ICU-level care and death according to age and asymptomatic percentage.

As the infection fatality rate (IFR) is a variable of great uncertainty, we include it in the model with a coefficient to mitigate its inaccuracy. As discussed below, an unknown lethality will not significantly affect the main objective of our study. Some epidemiological models of the late 2000s also failed to predict the evolution of H1N1 for the same reason—that it is necessary to know variables that are still uncertain at the beginning of an epidemic [46, 47]. Our *a priori* estimates of the IFR vary from 25% · *Prob_ICU_*, when the health system is not collapsed, to 100% · *Prob_ICU_*, when the health system collapsed, where *Prob_ICU_* is the probability that an infected individual require ICU-level care. We computed the probability of an individual requiring ICU-level care by using formula (1). The result of this formula to different regions can be seen in Table 1. For instance, for São Paulo, the probability that an infected individual will require ICU-level care is taken to be 0.6627%, whereas the median posterior empirical IFR was 0.36% (which is due to the fact that the model does not expect São Paulo’s health system to be collapsed). The *a posteriori* IFR varied from 55% · *Prob_ICU_*, when the health system is not collapsed, to 100% · *Prob_ICU_*. Thus, the model estimates the probability of death of an individual that requires ICU-level care to be from 55% to 100%. Our resulting IFR are very close to the ones obtained by using the figures from [48] and the corresponding age pyramid, which indicates that the magnitude of that IFR we used are likely to be correct. To compute the empirical estimate of the IFR, we suggest the reader to compute the estimated number of deaths at the end of august and divide to the number of infected individuals at the end of august. The reason is that if one takes the number of infected individuals, for instance in April, several individuals that will die and still have not died will appear in the denominator, whereas the corresponding death does not appear on the numerator. COVID-19’s IFR increases exponentially with age [8, 32, 23, 31]. The average age of the deaths in Italy was 80 years, for example [49]. So, considering our age pyramid with a much larger base than European and North American pyramids.

The mathematical model used in our study is based on an adaptive Bayesian technique that is based on the death toll, which we believe is the most reliable data we currently have in Brazil. The model has very different characteristics compared to other models previously used, as we use it oppositely from other models by providing the number of deaths as input. Considering symptomatic individuals, we know their chances of hospitalization, ICU treatment, and death [32, 23, 25]. Inexact knowledge of IFR will not change the outcome of the main objective of our study: to estimate the number of ICU beds needed over time. We consider this the most important variable in terms of public health, as it is possible to control with social isolation in order to save lives.

One possible criticism would be that we did not limit the study to just achieving herd immunity. It is not our goal to try to predict the total number of deaths at the end of the epidemic, but to help predict how many ICU beds would be needed over time. After the peak of deaths, we can see that there is a sharp drop in the curve. When the *R_t_* is less than 1 (that is, the time when 1 person infects less than 1 person), the curve no longer grows and the epidemic is controlled naturally. From that point onward, there is a marked decrease in the number of infected. As our population is in social isolation, the acute control of the epidemic would take place in this scenario. When the scenario changes, a further increase in the number of deaths is obviously expected. The magnitude of this increase will depend on how effective isolation is in controlling the disease. After the death rate peaks and there is a gradual loosening of isolation, we can envision either an optimistic scenario, in which the number of asymptomatic patients is higher than expected and the epidemic curve will tend to a more “endemic” characteristic (if the lethality is even lower than expected), or a pessimistic scenario, with a new peak of infection and deaths. At the end of this first stage of the epidemic, we estimated the number of deaths to be about 10 times lower than that from Imperial College’s publication [50]. Compartmental models are more sensitive to IFR than our model and can have the effect of overestimating the result. Another premise established by Ferguson et al. [34] is contact transmission. New evidence suggests transmission through the air without the need for contact [51].

For public health purposes, we consider that the lethality of the disease in a person who needs ICU care but does not receive it is 100%. As the average ICU stay is two weeks [23], ICU beds can quickly become unavailable. When we note that the peak need estimate for ICU beds is between late April and early May, after that peak (and if the number of deaths remains within our confidence interval), we could loosen the need for social isolation with well-established criteria and monitor whether more deaths are predicted beyond the capacity of ICU beds. By feeding our evolutionary curve with real data, we are able to monitor the epidemic’s evolution in real time. The daily routine has shown that our proposed model should behave in a more consistent manner with reality, bringing more realistic numbers to our country. We therefore offer an instrument that helps to predict the peak of deaths from the disease and, consequently, the need for ICU beds. According to the current scenario in terms of population isolation, there should be enough ICU beds in the state of São Paulo to face the epidemic. The situation may be more fearful, however, in other parts of the country where social inequality and the difference in health structure observed in Brazil would present a problem in facing the epidemic. We adapted our results according to these regions and will offer technical support to government officials according to different scenarios.

As the new corona virus dictates pandemics progression, probabilistic models should not be viewed as a static scientific truth but rather as an adaptative tool that may be useful to guide short term decisions in health and social policies when combined to a broader epidemiological context.

## Data Availability

Data will be available on demand.

